# AI-Powered Pipeline for Annotating Echocardiography Notes and Prognostic Variable Analysis in Critical Care

**DOI:** 10.64898/2026.03.09.26347835

**Authors:** Sheng Xu, Tammy Ma, Cindy Duan, April Ip, Calvin Tam, Yuki Yiu Ki Leung, Jiannan Yang, Simon Wai-Ching Sin, Emmanuel Cheung, Kai Hang Yiu, Pauline Yeung Ng

## Abstract

**Background:** Echocardiography (echo) notes contain valuable prognostic information for patients in the intensive care unit (ICU). However, their unstructured format and the presence of sensitive patient information present challenges for large-scale, automated analysis. There is a need for secure and efficient methods to extract and utilize echo data to enhance ICU outcome prediction.

**Methods:** We developed an AI-powered, privacy-preserving pipeline that leverages advanced natural language processing and pattern matching to annotate echo notes locally, ensuring comprehensive masking of personally identifiable information. This pipeline was applied to patient data from a mixed medical surgical ICU in a tertiary referral hospital. Key variables were extracted from echo notes and integrated with clinical and laboratory data to predict ICU mortality. A LightGBM machine learning model—robust to missing values—was trained using both routine and echo-derived structured clinical features. Its predictive performance was compared to that of the APACHE IV score.

**Results:** Compared with the reference standard derived from manual annotation by echocardiography specialist, automated annotation of echo notes achieved 98.85% data accuracy with a false positive rate of 0.31%. Several echo-derived variables, including left ventricular ejection fraction (LVEF), left ventricular outflow tract velocity time integral (LVOT VTI), tricuspid annular plane systolic excursion (TAPSE), mitral regurgitation (MR), and aortic regurgitation (AR), were strongly associated with ICU mortality. Incorporating echo-derived variables improved the accuracy in prediction of ICU mortality, with the LightGBM model achieving an AUC of 0.902 compared to 0.861 for APACHE IV score.

**Conclusion:** Our locally deployable AI pipeline enables secure and automated extraction of prognostic information from echo notes, substantially enhancing ICU mortality prediction. The inclusion of echo-derived variables significantly improved predictive accuracy, underscoring the potential but currently underutilized value of unstructured notes. This approach paves the way for scalable, privacy-preserving decision support tools in critical care.

## Introduction

The intensive care unit (ICU) is a highly complex environment in which accurate prediction of patient outcomes is crucial for effective clinical decision-making and optimal resource allocation. Widely-used mortality prediction scoring systems, such as SAPS II^1^ and APACHE IV^2^, have facilitated risk stratification and guided interventions; however, these models often lack the integration of nuanced diagnostic information available during the processes of patient care. Echocardiography (echo) is a cornerstone diagnostic modality in the ICU, providing essential insights into cardiac function and hemodynamic status — factors that are closely linked to patient prognosis^3^. Despite its clinical significance, the majority of echo data is embedded within unstructured clinical notes, complicating systematic extraction and incorporation into risk models. Several key variables documented in echo reports — such as left ventricular ejection fraction^4^ (LVEF), tricuspid annular plane systolic excursion^5^ (TAPSE), mitral regurgitation^6^ (MR), and tricuspid regurgitation^7^ (TR) — have been shown to be strongly associated with ICU mortality. However, because these parameters are typically recorded in free-text format in echo notes, they are not readily amenable to direct analysis or inclusion in prediction models.

Recent advances in natural language processing (NLP), particularly the development of large language models (LLMs), have created new opportunities for extracting meaningful information from unstructured clinical texts^8^. These models enable automated annotation and standardization of echo reports, thus facilitating integration with structured ICU datasets. Combining LLM-annotated echo data with comprehensive clinical variables holds the potential to improve the understanding of factors influencing ICU outcomes and to enhance the performance of predictive models^9^. Nevertheless, the use of online LLMs, such as OpenAI’s GPT-4 and Google’s Gemini, introduces challenges related to data privacy, security, and regulatory compliance. These models process data on remote servers, raising concerns about the handling of sensitive information, which could inadvertently be retained or used for model improvement despite stated privacy policies^10^. Although AI-enabled echo interpretation systems have been developed^11^, important limitations remain, including limited compatibility across different echo machine manufacturers and additional implementation costs.

In this study, we present a framework that leverages LLM-powered annotation of echo notes in conjunction with structured ICU clinical data to maximize inclusion of clinical information associated with ICU mortality, while ensuring data security through the exclusive use of a local LLM database with no external internet requirements. Focusing on patients from a mixed medical and surgical ICU, we jointly analyze echo reports and detailed clinical data to develop and evaluate predictive models for ICU mortality.

## Methods

### Study Population

This study included all patients admitted to the Intensive Care Unit (ICU) of Queen Mary Hospital, Hong Kong between October 2013 and April 2025, who underwent echocardiography (echo) examinations and had their echo reports recorded in the Clinical Information System (CIS). Echo reports and comprehensive ICU clinical data were extracted from a territory-wide electronic health record system and the ICU-specific CIS. Clinical data comprised patient demographic information, such as age and gender, as well as physiological variables and laboratory test results from the first 24 hours of ICU admission. These results included mean blood pressure (mean_BP), sodium (Na), potassium (K), hemoglobin (Hb), hematocrit (Hct), platelet count (Plt), and glucose (Glu), among others; for these variables, only the highest and lowest values within the first 24 hours were recorded. Blood gas measurements — including pH, bicarbonate (HCO_₃_), and fraction of inspired oxygen (FiO_₂_) — were treated as time-series data and recorded sequentially as first, second, and subsequent readings. Detailed clinical data and variables are provided in **Supplementary Table 1**.

Patients with an ICU stay of less than 4 hours or with missing APACHE IV scores were excluded from the dataset. For patients who did not survive the first 24 hours, only data recorded prior to the time of death were included. For each hospital admission, the first echo report was collected and analyzed. Patients with multiple ICU admissions (readmissions) were included, and all analyses were conducted on a per-admission basis.

### De-identification of echo Notes

Prior to data extraction and analysis, all echo notes were de-identified to protect potentially sensitive information. An English transformer-based model, **en_core_web_trf** in spaCy, was used to mask all English names as well as dates and times^12,13^. Additionally, a fine-tuned BERT model, **bert-base-NER**, was employed to mask Chinese pinyin names^14^.

### Transforming echo Notes into Structured Data

The de-identified echo notes were transformed into structured data using a combination of traditional pattern matching techniques and large language model (LLM) processing. To prioritize variables with high prevalence, we applied Term Frequency-Inverse Document Frequency (TF-IDF^15^) to all echo notes and selected clinically meaningful terms with high scores.

For this study, we identified a total of nine echo variables: Left Ventricular Ejection Fraction (LVEF), Inferior Vena Cava diameter (IVC), Left Ventricular Outflow Tract Velocity Time Integral (LVOT VTI), mitral valve peak E velocity to mitral annulus e’ velocity ratio (E/e’), Tricuspid Annular Plane Systolic Excursion (TAPSE), Pericardial Effusion (PE), Mitral Regurgitation (MR), Aortic Regurgitation (AR), and Tricuspid Regurgitation (TR). Of these, the first 5 variables (LVEF, IVC, LVOT VTI, E/e’, TAPSE) are continuous variables, while the remaining 4 (PE, MR, AR, TR) are categorical variables.

Extraction for LVEF, IVC, LVOT VTI, E/e’, TAPSE, and PE was performed using traditional pattern matching: relevant keywords were identified, and corresponding values were then extracted. For Pericardial Effusion (PE), after keyword identification, the extracted labels were compared against a predefined set of “no indicators” to determine the final classification as either “Yes” or “No”. LVEF can be measured using various methods, such as the modified Simpson’s biplane method, M mode, or by eyeballing. If no specific method was indicated, LVEF will be automatically recorded; if multiple measurements are available, priority was given to the modified Simpson’s biplane method.

MR, AR, and TR are often described in various styles within echo notes; therefore, their annotation was performed using LLM. Specifically, the google/gemma-3-12b model was downloaded and implemented via LM Studio to assist in annotating these variables^16^. Severity for each valvular regurgitation was categorized as: no, trivial, mild, moderate, or severe. If the model could not identify a clear severity annotation, the variable was labeled as “detected”. In cases where no relevant keywords for MR, AR, or TR were found, the annotation was set to “unknown”. To mitigate potential false-positive annotations from the LLM, we verified outputs using rule-based pattern matching on the echo notes. When an exact variable name could not be identified in the notes, the annotation was set to “unknown.”

### Validation of Annotation Results

The final annotation results were validated against a reference standard, defined as manual annotation by a certified echocardiography specialist. A randomly selected subset of echo notes was annotated and served as the reference standard. Only the following parameters were included in the manual annotation: LVEF, PE, IVC, MR, AR, and TR. These parameters were used to evaluate and compare the accuracy of the automated annotations.

Definitions are as follows:

False negative: a parameter value present in the echo notes but not detected by the automated method.

Correct: the detected value matches the reference standard. Incorrect: the detected value does not match the reference standard.

False positive: a value output for a parameter that has no corresponding value in the echo notes.

The evaluation of the annotation results was based on three criteria:

1. Data completeness: the proportion of parameters present in the echo notes that were captured by the automated method, regardless of correctness. Formula: (Correct + Incorrect) / (False negative + Correct + Incorrect).
2. Data accuracy: the proportion of captured parameters that matched the reference standard. Formula: Correct / (Correct + Incorrect).
3. Data false positive rate: the proportion of parameters output by the automated method that were not present in the echo notes. Formula: False Positives / (Correct + Incorrect + False Positives + False negative).

### Statistical Methods

The Wilcoxon rank sum test (Mann-Whitney U test) was used to compare individual continuous variables with the primary study outcome – ICU mortality. P values were adjusted for multiple testing using the false discovery rate (FDR) method. For categorical variables, the chi-square test was performed, with FDR adjustment applied to the resulting p values. To minimize false positives, a stringent threshold for significance was set at an adjusted p value < 0.01.

### Dimensional Reduction and Overall Difference Comparison

Dimensionality reduction and data exploration were performed using PCAmix (Principal Component Analysis of Mixed Data) from the R package “PCAmixdata”^17^. PCAmix was applied to all complete cases (excluding observations with missing values), reducing the data to five principal components. To assess overall differences in the reduced data, PERMANOVA (Permutational Multivariate Analysis of Variance) was conducted using the R package “vegan”^18^. The analysis employed 999 permutations and utilized the “euclidean” distance metric.

### Bayesian Network

Bayesian networks were utilized to explore the conditional dependencies between echo or clinical variables and ICU mortality, using the R package “bnlearn”^19^. The Hill Climbing (HC) algorithm was implemented to determine the optimal network structure within the Bayesian framework. ICU mortality was analyzed as the outcome variable and designated as a leaf node in the network.

### Linear Mixed-Effects Model

Linear mixed-effects models were fitted using the R packages “lme4” and “lmerTest” to analyze the longitudinal data^20,21^. These models accounted for both the main effects and the interaction between ICU mortality status and time, as well as the correlation of repeated measures within individuals. A random intercept for each patient was included to account for within-subject variability. The model formula was specified as:

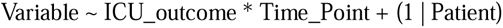

Where “Variable” represents the dependent variable of interest (e.g., pH, HCO_3_^-^, or FiO_2_), the asterisk (*) denotes the interaction effect between ICU outcome and time point, and “(1 | Patient)” represents the random intercept for each patient.

### Prediction of ICU Mortality with Machine Learning Approach

The dataset was split into training and testing cohorts based on the timing of the echo examinations. All cases with echo exams conducted after 2024 were assigned to the testing cohort, while the remaining cases comprised the training cohort.

Feature selection was conducted using two approaches. For categorical variables, a random forest algorithm (R package: “randomForest”^22^) was applied, and variables with a variable importance greater than 10 were retained. For continuous variables, Wilcoxon rank-sum tests were performed to compare values between survivors and non-survivors; variables with a p-value less than 0.01 were selected. Highly correlated features (Pearson correlation coefficient > 0.7) were excluded to minimize multicollinearity.

Two machine learning algorithms were used to predict ICU mortality: LightGBM (R package: “lightgbm”^23^) and logistic regression. Model hyperparameters—including number of leaves, learning rate, and feature fraction—were optimized using Bayesian optimization (R package: “ParBayesianOptimization”^24^) with three-fold cross-validation.

Three models were developed for comparison:

Model 1: LightGBM trained on all selected features.
Model 2: LightGBM trained on all selected features except those derived from echo notes.
Model test (Comparator): Logistic regression trained solely on the Apache IV score.

Model performance was evaluated using the area under the curve (AUC) to compare predictive accuracy across the three approaches.

### Risk score derivation and probability estimation

Risk scores (logits) were generated using Model 1. For each individual i, let s_i denote the logit score. The probability of death (Risk_i) was estimated as follows:

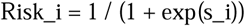

In this formulation, a higher risk score indicates a lower probability of death, while a lower corresponds to a higher probability of death.

## Results

### Study Population and Characteristics

The study design is illustrated in **Figure 1**. A total of 4,632 echo examinations were collected. Of these, 4,377 (94.5%) had corresponding clinical data and echo notes that documented at least one of the nine selected echo variables. Since each patient could undergo multiple echo tests during a single hospital admission, we limited our downstream analysis to only the first echo note per patient per hospital admission to avoid bias from repeated measures. This resulted in a final dataset of 2,976 echo notes from 2,755 patients, including 189 with multiple notes from different ICU admissions. There was a total of 1,154 females (38.8%), with a median age of 65 years and a standard deviation of 15 years. Key clinical and ECHO variables are presented in **Table 1**.

**Figure 1.**
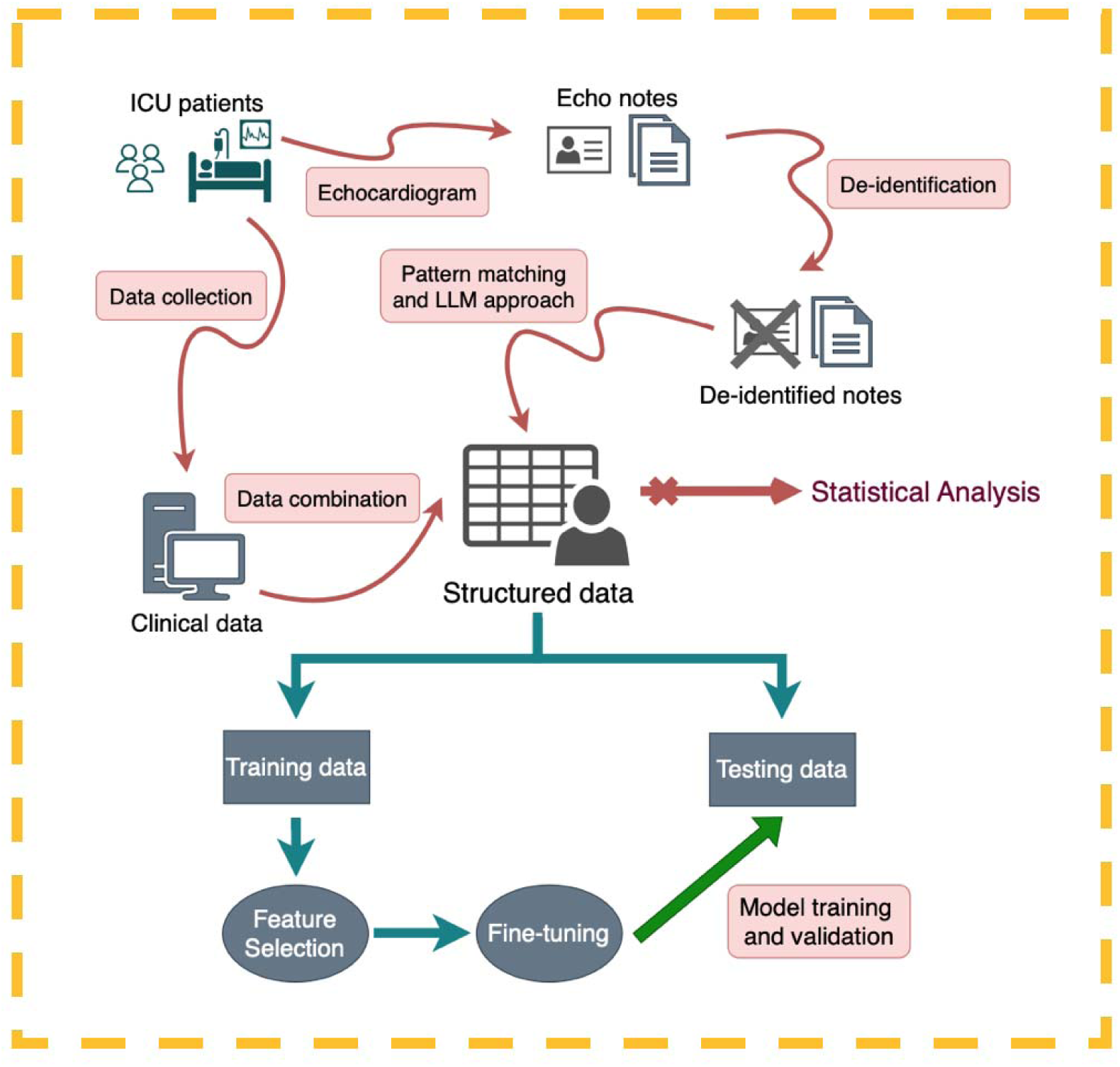
Overview of the Study Workflow

**Table 1:** Key Clinical and Echocardiographic Variables.

| Characteristic | Died<br>N = 581<sup>1</sup> | Alive<br>N = 2,395<sup>1</sup> | p-value<sup>2</sup> | Model-selected<sup>3</sup> |
| --- | --- | --- | --- | --- |
| Age (year) | 65 (15), N = 581 | 65 (15), N = 2395 | 0.3 | N |
| Gender |  |  | 0.3 | Y |
| Female | 213 / 580 (37%) | 941 / 2,391 (39%) |  |  |
| LVEF (%) | 43 (18), N = 431 | 48 (16), N = 1780 | <0.001 | Y |
| IVC diameter (cm) | 1.60 (0.52), N = 308 | 1.64 (0.57), N = 1382 | 0.4 | N |
| LVOT VTI (cm) | 14.7 (6.1), N = 157 | 16.5 (5.6), N = 900 | <0.001 | Y |
| E/e' | 11.0 (4.4), N = 146 | 10.9 (5.1), N = 842 | 0.3 | N |
| TAPSE (cm) | 1.70 (0.55), N = 121 | 1.82 (0.49), N = 672 | 0.018 | Y |
| Apache IV score | 130 (37), N = 581 | 86 (28), N = 2395 | <0.001 | Y |
| Highest core body temperature | 37.34 (1.31), N = 579 | 37.76 (1.00), N = 2389 | <0.001 | Y |
| Lowest Core body temperature | 35.59 (1.45), N = 581 | 36.23 (0.99), N = 2389 | <0.001 | Y |
| Highest heart rate per minute | 123 (25), N = 581 | 116 (24), N = 2395 | <0.001 | Y |
| Lowest heart rate per minute | 76 (22), N = 560 | 78 (18), N = 2395 | 0.3 | N |
| Highest respiratory rate per minute | 33 (8), N = 581 | 31 (8), N = 2395 | <0.001 | Y |
| Lowest respiratory rate per minute | 14.1 (4.2), N = 560 | 13.1 (3.6), N = 2395 | <0.001 | Y |
| Highest mean blood pressure (mmHg) | 96 (18), N = 581 | 103 (20), N = 2395 | <0.001 | Y |
| Lowest mean blood pressure (mmHg) | 50 (14), N = 561 | 60 (12), N = 2395 | <0.001 | Y |
| 24 hours urine (ml) | 870 (1,169), N = 581 | 1,463 (1,177), N = 2395 | <0.001 | Y |
| Highest creatinine (umol / L) | 296 (219), N = 580 | 259 (281), N = 2392 | <0.001 | Y |
| Highest bilirubin (umol / L) | 84 (148), N = 580 | 38 (74), N = 2392 | <0.001 | Y |
| Highest WCC (10<sup>-9</sup> / L) | 18 (35), N = 580 | 16 (14), N = 2392 | 0.4 | N |
| 1<sup>st</sup> pH | 7.26 (0.18), N = 581 | 7.33 (0.14), N = 2391 | <0.001 | Y |
| 1<sup>st</sup> PaCO<sub>2</sub> (kPa) | 5.07 (2.29), N = 574 | 5.12 (2.35), N = 2376 | 0.3 | N |
| 1<sup>st</sup> PaO<sub>2</sub> (kPa) | 20 (17), N = 574 | 17 (12), N = 2376 | 0.13 | N |
| 1<sup>st</sup> HCO<sub>3</sub> | 17 (7), N = 574 | 20 (7), N = 2376 | <0.001 | Y |
| 1<sup>st</sup> FiO<sub>2</sub> | 0.62 (0.29), N = 581 | 0.50 (0.28), N = 2391 | <0.001 | Y |
| Pericardial effusion |  |  | 0.3 | Y |
| No | 360 / 581 (62%) | 1,545 / 2,395 (65%) |  |  |
| Yes | 74 / 581 (13%) | 252 / 2,395 (11%) |  |  |
| MR |  |  | <0.001 | Y |
| detected<sup>4</sup> | 1 / 581 (0.2%) | 9 / 2,395 (0.4%) |  |  |
| mild | 89 / 581 (15%) | 493 / 2,395 (21%) |  |  |
| moderate | 46 / 581 (7.9%) | 164 / 2,395 (6.8%) |  |  |
| no | 63 / 581 (11%) | 366 / 2,395 (15%) |  |  |
| severe | 5 / 581 (0.9%) | 31 / 2,395 (1.3%) |  |  |
| trivial | 29 / 581 (5.0%) | 139 / 2,395 (5.8%) |  |  |
| AR |  |  | <0.001 | Y |
| detected<sup>4</sup> | 3 / 581 (0.5%) | 10 / 2,395 (0.4%) |  |  |
| mild | 40 / 581 (6.9%) | 206 / 2,395 (8.6%) |  |  |
| moderate | 19 / 581 (3.3%) | 64 / 2,395 (2.7%) |  |  |
| no | 59 / 581 (10%) | 406 / 2,395 (17%) |  |  |
| severe | 1 / 581 (0.2%) | 12 / 2,395 (0.5%) |  |  |
| trivial | 13 / 581 (2.2%) | 88 / 2,395 (3.7%) |  |  |
| TR |  |  | 0.068 | Y |
| detected<sup>4</sup> | 21 / 581 (3.6%) | 72 / 2,395 (3.0%) |  |  |
| mild | 90 / 581 (15%) | 392 / 2,395 (16%) |  |  |
| moderate | 47 / 581 (8.1%) | 240 / 2,395 (10%) |  |  |
| no | 41 / 581 (7.1%) | 218 / 2,395 (9.1%) |  |  |
| severe | 18 / 581 (3.1%) | 59 / 2,395 (2.5%) |  |  |
| trivial | 27 / 581 (4.6%) | 162 / 2,395 (6.8%) |  |  |
| ICU Admission type |  |  | <0.001 | N |
| Elective | 4 / 581 (0.7%) | 116 / 2,395 (4.8%) |  |  |
| Emergency | 577 / 581 (99%) | 2,279 / 2,395 (95%) |  |  |
| ICU admission source |  |  | <0.001 | Y |
| A&E | 102 / 581 (18%) | 422 / 2,395 (18%) |  |  |
| General ward | 303 / 581 (52%) | 1,147 / 2,395 (48%) |  |  |
| OT/Recovery | 50 / 581 (8.6%) | 470 / 2,395 (20%) |  |  |
| Others | 126 / 581 (22%) | 356 / 2395 (15%) |  |  |
| Chronic health |  |  | <0.001 | Y |
| Yes | 302 / 581 (52%) | 1,049 / 2,395 (44%) |  |  |
| Ventilator support during ICU stay (Yes) | 174 / 265 (66%) | 427 / 1,379 (31%) | <0.001 | N |
| Renal replacement (Yes) | 315 / 565 (56%) | 510 / 2,367 (22%) | <0.001 | Y |
| 1<sup>st</sup> Endotracheal tube or tracheostomy |  |  | <0.001 | N |
| N | 335 / 581 (58%) | 1,583 / 2,391 (66%) |  |  |
| Y | 246 / 581 (42%) | 808 / 2,391 (34%) |  |  |
| 1<sup>st</sup> Mechanical ventilation |  |  | <0.001 | N |
| N | 310 / 581 (53%) | 1,527 / 2,391 (64%) |  |  |
| Y | 271 / 581 (47%) | 864 / 2,391 (36%) |  |  |
| 1<sup>st</sup> Arterial Vensus |  |  | 0.8 | N |
| A | 553 / 581 (95%) | 2,285 / 2,391 (96%) |  |  |
| V | 28 / 581 (4.8%) | 106 / 2,391 (4.4%) |  |  |

<sup>1</sup>Mean (SD); n / N (%)
<sup>2</sup>Wilcoxon rank sum test; Pearson's Chi-squared test; Wilcoxon rank sum exact test; NA
<sup>3</sup> Indicate whether the variable was selected for predicting ICU mortality in the later section. (Y/N)
<sup>4</sup> Detected: MR/AR/TR was identified, however, a clear severity annotation could not be determined
\*\*LVEF: Left Ventricular Ejection Fraction; IVC: Inferior Vena Cava; LVOT VTI: Left Ventricular Outflow Tract Velocity Time Integral; E/e': mitral valve peak E velocity to mitral annulus e' velocity ratio; TAPSE: Tricuspid Annular Plane Systolic Excursion; MR: Mitral Regurgitation; AR: Aortic Regurgitation; TR: Tricuspid Regurgitation.

### Accuracy of Automated Annotation of echo Notes

We annotated echo notes using both pattern matching and LLMs, and compared the results to the reference standard derived by manual annotation. Both approaches achieved high accuracy, high completeness, and low false positive rates, indicating excellent overall performance (**Table 2**). The overall automated annotation achieved an accuracy of 98.85%, with a completeness of 98.97% and a false positive rate of 0.31%. Performance varied across different variables: for example, both LVEF and IVC reached an accuracy of 100%, while TR had a lower accuracy of 95.45%. False positives were highly controlled throughout the process; only one false positive was identified in IVC, and none were observed for the other variables.

**Table 2.** Accuracy, Completeness, and False-Positive Rates for Automated Annotation of Echo Notes Compared with Reference Standard^1^.

| Variable | Detection Method | Observations<br>(Non-NA) | Data<br>Completeness<sup>2</sup> | Data Accuracy<sup>3</sup> | Number of False Positives<br>(False positive rate)<sup>4</sup> |
| --- | --- | --- | --- | --- | --- |
| LVEF | Pattern matching | 81 | 80/81 (98.77%) | 80/80 (100%) | 0 (0%) |
| Pericardial Effusion | Pattern matching | 86 | 85/86 (98.84%) | 85/85 (100%) | 0 (0%) |
| IVC diameter | Pattern matching | 53 | 51/53 (96.23%) | 51/51 (100%) | 1 (1.85%) |
| MR | Large language model | 42 | 42/42 (100%) | 41/42 (97.62%) | 0 (0%) |
| AR | Large language model | 21 | 21/21 (100%) | 21/21 (100%) | 0 (0%) |
| TR | Large language model | 44 | 44/44 (100%) | 42/44 (95.45%) | 0 (%) |
| Overall<sup>5</sup> |  |  | 98.97% | 98.85% | 0.31% |

1. In total 101 echo notes.
2. Data completeness: the proportion of parameters present in the echo notes that were captured by the automated method, regardless of correctness
3. Data accuracy: the proportion of captured parameters that matched the reference standard.
4. Data false positive rate: the proportion of parameters output by the automated method that were not present in the echo notes.
5. Overall: average performance across variables.
\*\*LVEF: Left Ventricular Ejection Fraction; IVC: Inferior Vena Cava; MR: Mitral Regurgitation; AR: Aortic Regurgitation; TR: Tricuspid Regurgitation.

Some errors arose from unexpected writing styles or mistakes within the raw echo notes, which posed challenges for both patterns matching and LLM-based methods. For example, one LVEF value was incorrectly annotated due to an unexpected comma between the variable name and its value (raw note: “LVEF fair, ∼40%”). Another error resulted from a typographical mistake in the original note, where “percardial effusion” was written instead of “pericardial effusion,” significantly impacting annotation accuracy.

### Association between echo Variables and ICU Mortality

The structured echo data were examined for association with ICU mortality (**Figure 2A** and **Supplementary Figure 1**). Among the nine variables examined, six variables (LVEF, VTI, TAPSE, MR, AR, and TR) were significantly associated with ICU mortality in univariable analysis. Specifically, the values of LVEF, VTI, and TAPSE were significantly higher in patients who survived compared to those who died (P < 0.001, P < 0.001, and P = 0.018, respectively). Additionally, the severity of MR, AR, and TR was also significantly related to ICU mortality; patients with “no,” “trivial,” or “mild” severity were more likely to survive.

**Figure 2.**
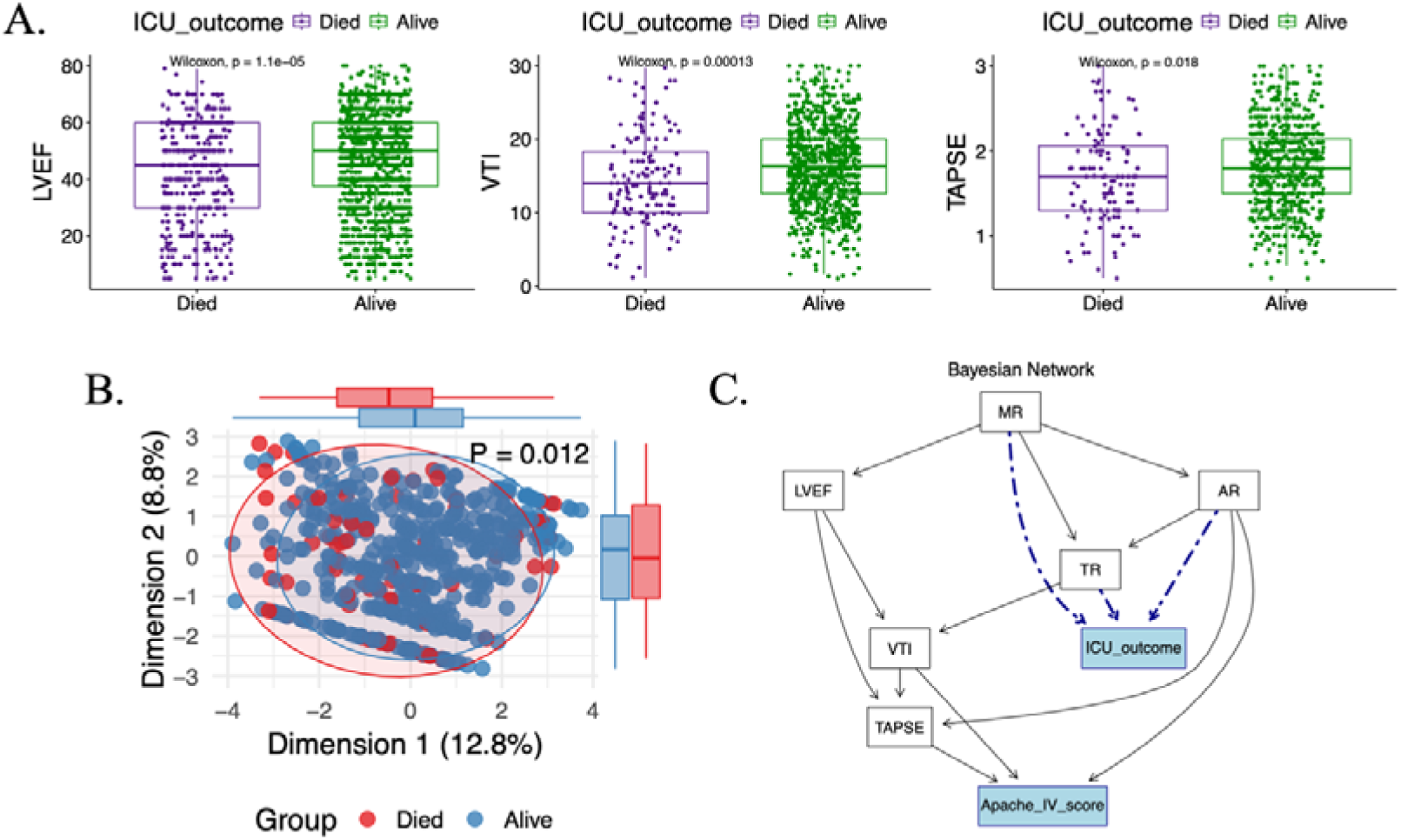
Associations Between Structured Echocardiographic Variables and ICU Mortality A. Associations of LVEF, VTI, and TAPSE with ICU mortality B. Principal component analysis of mixed data (PCAmix) plot illustrating differences by ICU mortality C. Causal relationships inferred from Bayesian network analysis

After univariable analyses, a comprehensive analysis including all the above 6 variables was conducted. Among the echo data, only 519 records contained complete information for all these variables; therefore, only these records were included in the subsequent analysis. A significant overall difference in these variables was observed between patients who survived ICU and those who did not, with a P value of 0.012 **(Figure 2B**). Causal relationships were estimated using a Bayesian Network, with the APACHE IV score included for comparison. The analysis showed that MR, AR, and TR were directly associated with ICU mortality, while the other variables were indirectly associated (**Figure 2C**).

### Association between clinical data and ICU Mortality

A total of 134 continuous variables were analyzed, including 39 independent variables and 5 variables (pH, PaCO_₂_, PaO_₂_, HCO_₃_, and FiO_₂_) with 19 time-series measurements each. Additionally, 69 categorical variables were assessed, comprising 12 independent variables and 3 variables with 19 time-series measurements each (endotracheal tube or tracheostomy, mechanical ventilation status, and arterial versus venous blood sampling). A total of 9 variables extracted from echo notes were included in the analysis.

Univariate analysis identified 24 continuous clinical variables significantly associated with ICU mortality (adjusted p < 0.01). APACHE IV scores were significantly higher in patients who died, alongside higher maximum heart rates, lower maximum and minimum blood pressures, lower maximum and minimum core temperatures, and higher maximum and minimum respiratory rates (**Figure 3**). Contrary to common assumptions, age was not associated with ICU mortality (p-value 0.34) (**Supplementary Figure 2A**).

**Figure 3.**
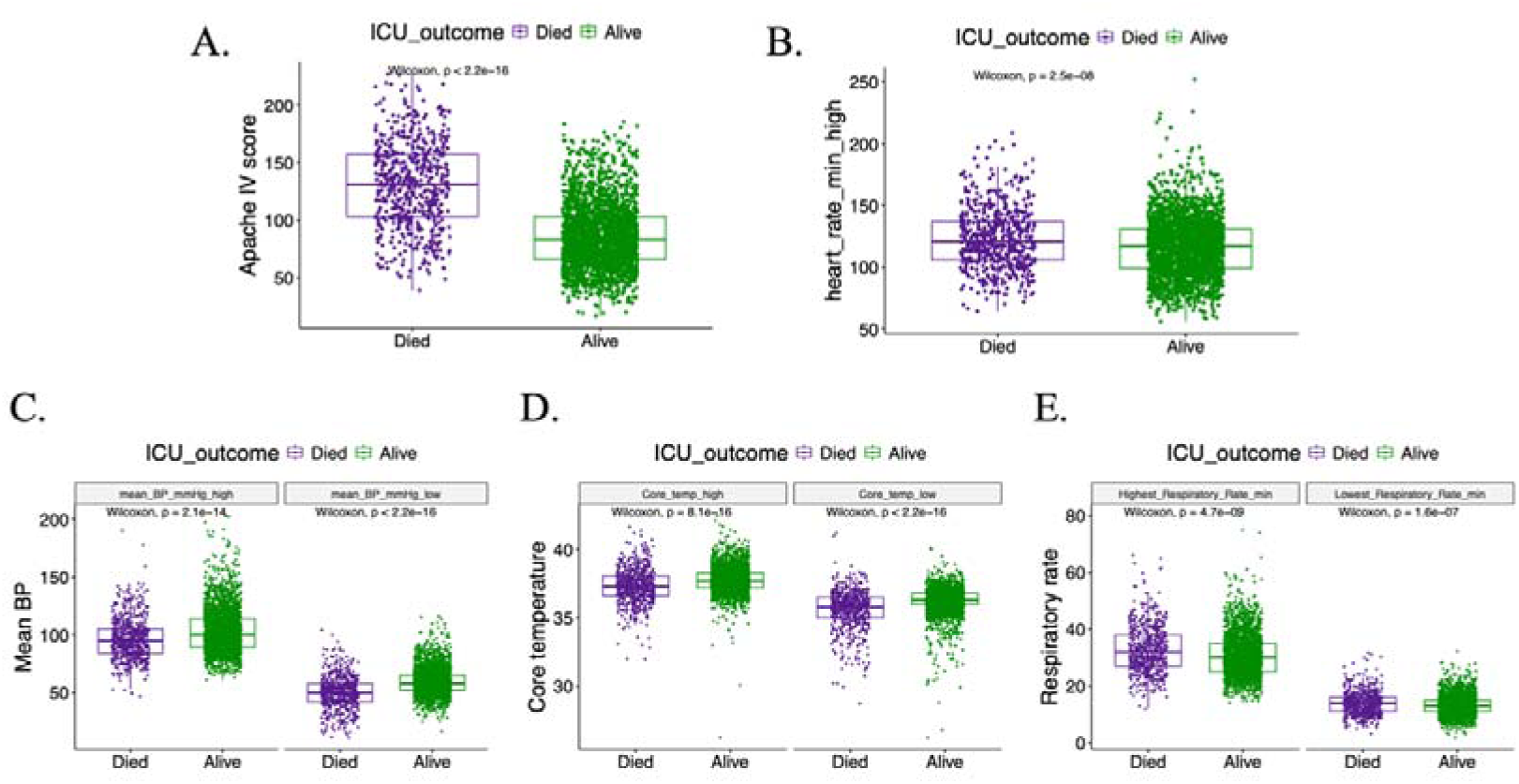
Associations Between Clinical Parameters and ICU Mortality A. APACHE IV score and ICU mortality B. Highest heart rate (beats per minute) and ICU mortality C. Highest and lowest mean blood pressure and ICU mortality D. Highest and lowest core temperature and ICU mortality E. Highest and lowest respiratory rate and ICU mortality

Among laboratory variables, several significant associations were observed, with some variables displaying high inter-correlation, particularly urea and creatinine (Creat), as well as hemoglobin (Hb) and hematocrit (Hct) (**Supplementary Figure 2B**). After removing highly correlated variables and retaining only one from each correlated pair, the remaining variables that were significantly associated with ICU mortality included Na, K, urea, glucose, Plt, Hb, albumin (Alb), bilirubin (Bil), and urine output (**Supplementary Figure 3**).

For variables measured repeatedly over time, levels of pH, HCO_3-_, and FiO_2_ were significantly associated with ICU mortality (**Supplementary Figure 4**). When each time point was analyzed independently, the first ten measurements for all three variables were significantly different between patients who died and those who survived. To jointly analyze data across all time points, a random intercept was included for each patient to account for the correlation of repeated measures within individuals. Using linear mixed-effects models, pH, HCO_3_-, and FiO_2_ were found to be significantly associated with both ICU mortality and time, with all p-values less than 0.001.

Significant differences were also observed in categorical variables, including chronic health status, the requirement for ventilator support, renal replacement, type of ICU admission, and source of admission (**Supplementary Figure 5**) between patients who died in ICU and those who did not. Gender did not differ significantly in our dataset.

### Prediction of ICU Mortality Using Clinical and Echocardiography Data

Clinical and structured echo data were integrated to predict ICU mortality in multivariable analysis. The final training cohort consisted of 2,317 cases, and the final testing cohort included 659 cases. Given the possibility of significant missing information in both clinical and echo notes, the prediction model was designed to tolerate missing values (NA). To address this, LightGBM was utilized, as it can effectively handle missing data without the need for imputation or removal. After feature selection and elimination of highly correlated variables, a total of 34 features were retained, including 10 categorical and 24 continuous variables (**Supplementary Table 2** and **Supplementary Figure 6**). Key variables extracted from echo notes included LVEF, VTI, TAPSE, PE, MR, AR, and TR.

Three machine learning models were developed and their performance compared using ROC curves. Model 1, which incorporated both clinical and echo data, achieved the highest performance compared with Model 2, which did not include echo data, and Model test (Comparator), which included only the APACHE IV score (AUC 0.902 vs 0.896 vs 0.861) (**Figure 4A**).

**Figure 4.**
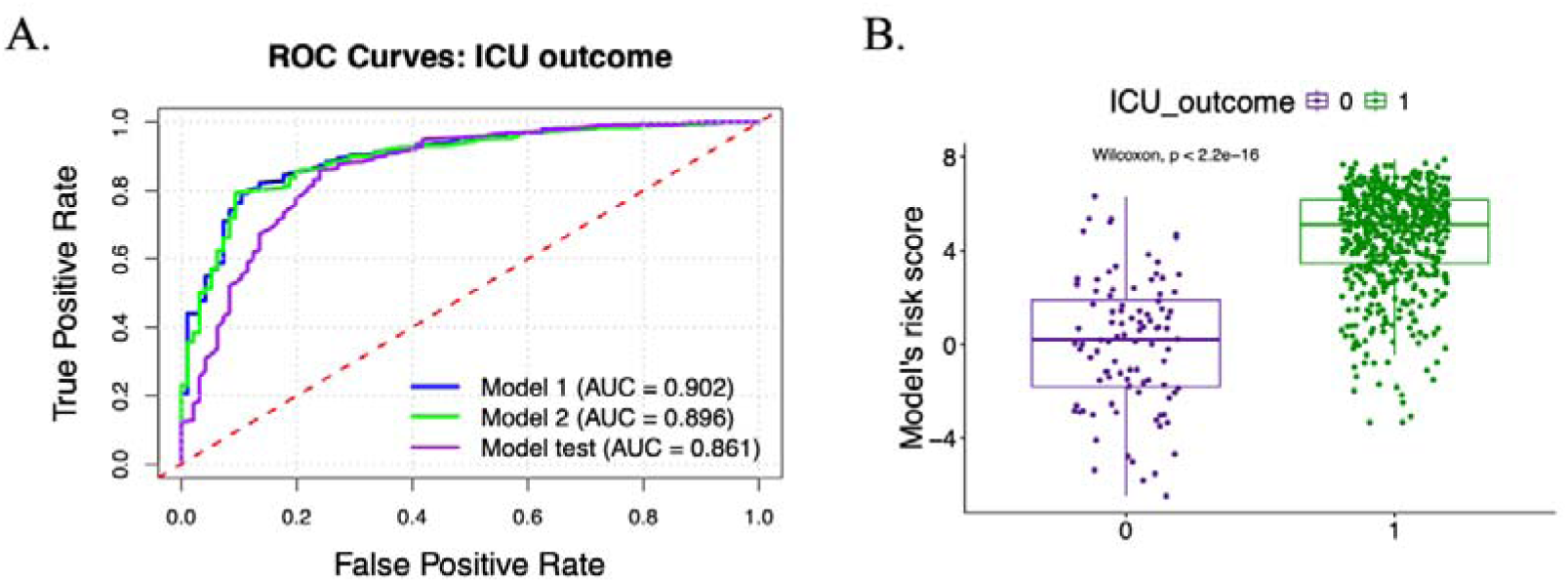
ICU Mortality Prediction Using echo Notes and Clinical Data A. Receiver operating characteristic (ROC) curves illustrating the performance of three models: Model 1: All features included; Model 2: All features except those derived from ECHO notes; Model 3: APACHE IV score only. B. Distribution of Model 1 risk scores in relation to ICU mortality outcomes in the testing cohort.

Risk scores were generated using Model 1 and compared between ICU patients who died and those who survived. Patients who died had significantly lower risk scores (median: 0.2), while survivors had higher scores (median: 5.1). This difference was statistically significant (P < 0.001; **Figure 4B**).

## Discussion

We introduced a locally-deployable AI-powered pipeline for annotating echo notes, combining advanced natural language processing techniques with traditional pattern matching to extract structured data from clinical reports. This approach provided important additional information to facilitate downstream clinical use, such as improving the accuracy of predicting ICU mortality compared to traditional risk scores. The entire pipeline executed efficiently, with minimal demands on computing infrastructure, enabling rapid annotation and analysis of echo notes with low risk of data leak.

Structured databases facilitate integration with other data types, enabling large-scale analyses and further advancement^25^. Traditional risk models in the ICU, such as APACHE IV, utilize discrete physiological and diagnostic data to predict ICU mortality risk^2^. However, unstructured data such as free text also hold significant value in the intensive care unit (ICU). For example, the incorporation of clinical notes has been shown to enhance early prediction of mortality and prolonged ICU stay^26^. Unstructured echocardiography (echo) reports contain rich clinical information about patients’ conditions, yet because echo parameters are not readily entered in structured case record forms, they are rarely included in studies involving ICU populations or prediction of outcomes.

Recent advances have introduced various methods for transforming unstructured text into structured data. Notably, large language models (e.g., ChatGPT) have been utilized to extract relevant information from medical notes. For instance, diagnostic details have been retrieved from unstructured lung cancer pathology notes to assist in developing treatment plans^27^. Nevertheless, these approaches raise critical concerns regarding data privacy as clinical information may have to be uploaded onto online servers. Additionally, the conversion of free text to structured formats often results in a high proportion of missing values, which presents a significant challenge for data analysis. Currently, most ICU studies rely on well-established scoring systems, such as the APACHE score, and utilize machine-learning algorithms that require complete data inputs. Methods like artificial neural networks^28^ and logistic regression^29^ are unable to accommodate missing values, further limiting the use of unstructured data in predictive modeling within the ICU setting.

Our automated annotation pipeline, combining rule-based pattern matching with LLMs, demonstrated near-perfect performance relative to the reference standard, achieving 98.85% data accuracy and a false positive rate of only 0.31%. When applied to echo notes from patients admitted to a mixed medical surgical ICU, the pipeline enabled a comprehensive analysis to identify variables potentially associated with ICU mortality. Several parameters extracted from echo notes—including LVEF, LVOT VTI, TAPSE, MR, and AR—demonstrated strong associations with mortality. LVEF, a widely recognized measure of cardiac function and a primary criterion for classifying heart failure^30^, was significantly lower in non-survivors, consistent with prior reports linking reduced LVEF to increased mortality risk^4^. Similarly, LVOT VTI, which reflects systolic function and cardiac output, was significantly lower in non-survivors^31^. TAPSE, an indicator of right ventricular systolic function, was also reduced in patients who died, aligning with evidence that lower TAPSE values are associated with higher mortality^32^. Although MR and AR are more challenging to quantify and are often recorded only in unstructured notes, they remain clinically important which may reflect their physiological association with fluid status and hemodynamics^6,33^. The ability of our pipeline to reliably capture these variables from free-text documentation prevents the loss of important clinical parameters to inform patients’ clinical condition and prognosis.

To balance privacy, practicality, and clinical utility, we implemented a stringent de-identification workflow that masked all personally identifiable information prior to analysis and deployed the entire pipeline on secure, on-premise systems. Given the sensitivity of clinical notes, we leveraged a local LLM pipeline so that all processing and storage remain confined to a protected, local environment. This design minimizes risks of data leakage, avoids reliance on external cloud services, and remains user-friendly with modest computational requirements, rendering it a practicable solution for real-world clinical adoption. For prediction, we combined variables extracted from echo notes with routinely collected ICU clinical data and trained a LightGBM model, selected for its native accommodation of missing values during both training and inference. This capability is particularly valuable in ICU settings, where random missingness is prevalent in both structured records and information derived from unstructured notes. In contrast, commonly used alternatives such as logistic regression and random forests typically require explicit imputation, which may be impractical or unreliable in deployment. An ablation analysis comparing models with and without echo-derived features showed that incorporating these variables significantly improved performance, highlighting the clinical relevance of echo documentation among ICU patients undergoing echo. The final model achieved an AUC of 0.902, substantially outperforming the widely used APACHE IV score, a strong benchmark for ICU prognostication^34^, and highlighting the added value of securely harnessing information from unstructured echo notes to better inform mortality risk stratification.

There are several limitations to our work. First, although the overall sample size of the number of echocardiography notes was reasonable, there was a relatively high proportion of missing values in the structured echo data. Secondly, our study is limited to a single unit, and it is possible that the AI-powered echo notes annotation pipeline we developed performs differently with echo notes from other hospitals. It would be useful to validate our approach with an external dataset. Third, our analysis included only patients who required echo, resulting in a refined cohort that may not be representative of the entire ICU population. Notably, age was not a significant factor in our dataset — at least among patients who required echo — despite age being a widely recognized predictor of ICU mortality^35^. Lastly, extracting more complex data, such as velocity measurements and indices of intracardiac pressures, may be even more challenging for an automated pipeline and require a larger sample size for training. Additionally, our ICU mortality prediction model was developed based on real-world application. Specifically, it was trained on older data and evaluated on relatively newer data, simulating the intended use of applying the model to future patient cases. However, this approach may introduce bias, as echocardiography techniques, staff expertise, and patient populations may have changed over time.

Overall, our findings highlight the value of incorporating structured information from echo notes into ICU prognostic models, and highlight the potential of AI-driven approaches to enhance clinical decision-making in critical care. The successful integration of AI methodologies demonstrates the feasibility of automating complex data extraction and analysis, paving the way for scalable, real-time decision support tools in the ICU. Ultimately, our study provided evidence for the continued adoption of data-driven technologies in critical care, with the aim of empowering clinicians and improving patient outcomes.

## Data Availability

All data produced in the present study are available upon reasonable request to the authors

## Declarations

## Ethical Approval and Consent to participate

This study was approved by the Institutional Review Board (IRB) of the University of Hong Kong / Hospital Authority Hong Kong West Cluster (IRB Reference Number: UW 24-434, dated 17th July 2024)

## Consent for publication

All authors consent for publication.

## Availability of supporting data

The datasets generated and/or analyzed during the proposed study will be available from the corresponding author upon reasonable request.

## Authors’ contributions

Sheng Xu conceived and designed the study, performed the experiments, analyzed the data, and drafted the manuscript. Pauline Yeung NG provided substantial modifications to the manuscript, supervised the research process, and managed correspondence with the journal. All the other authors contributed valuable suggestions and critical feedback. All the authors read and approved the final version of the manuscript.

## Acknowledgements

Not applicable.

---

**Supplementary Figure 1.**
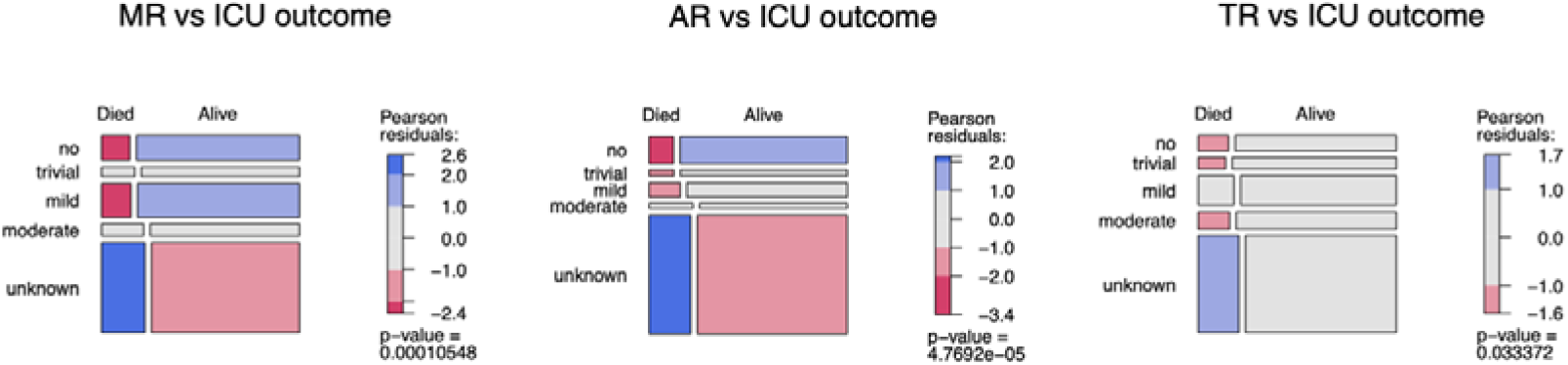
Mosaic Plot Depicting the Association of MR, AR, and TR with ICU Mortality Groups with insufficient observations were excluded from the figures, including the “detected” and “severe” patient groups.

**Supplementary Figure 2.**
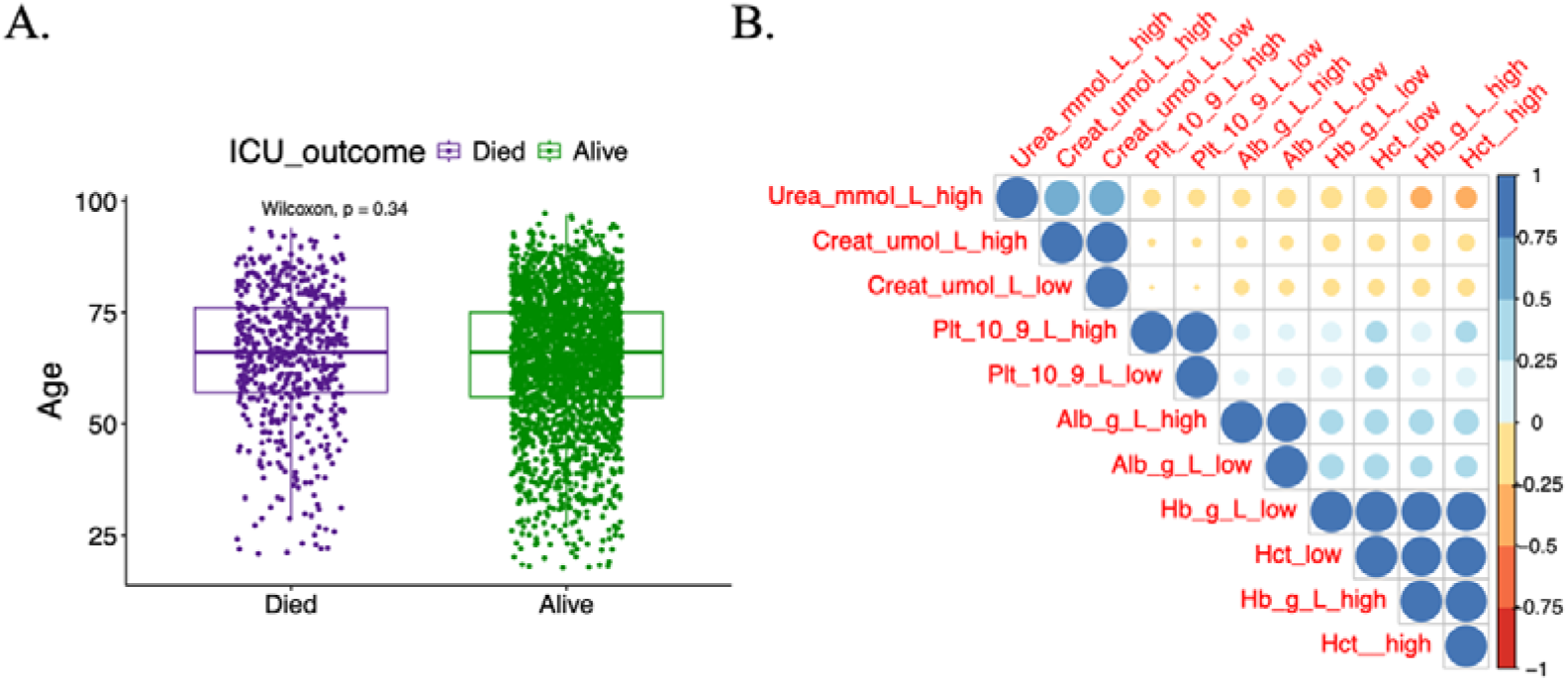
Age and other clinical variables

**Supplementary Figure 3.**
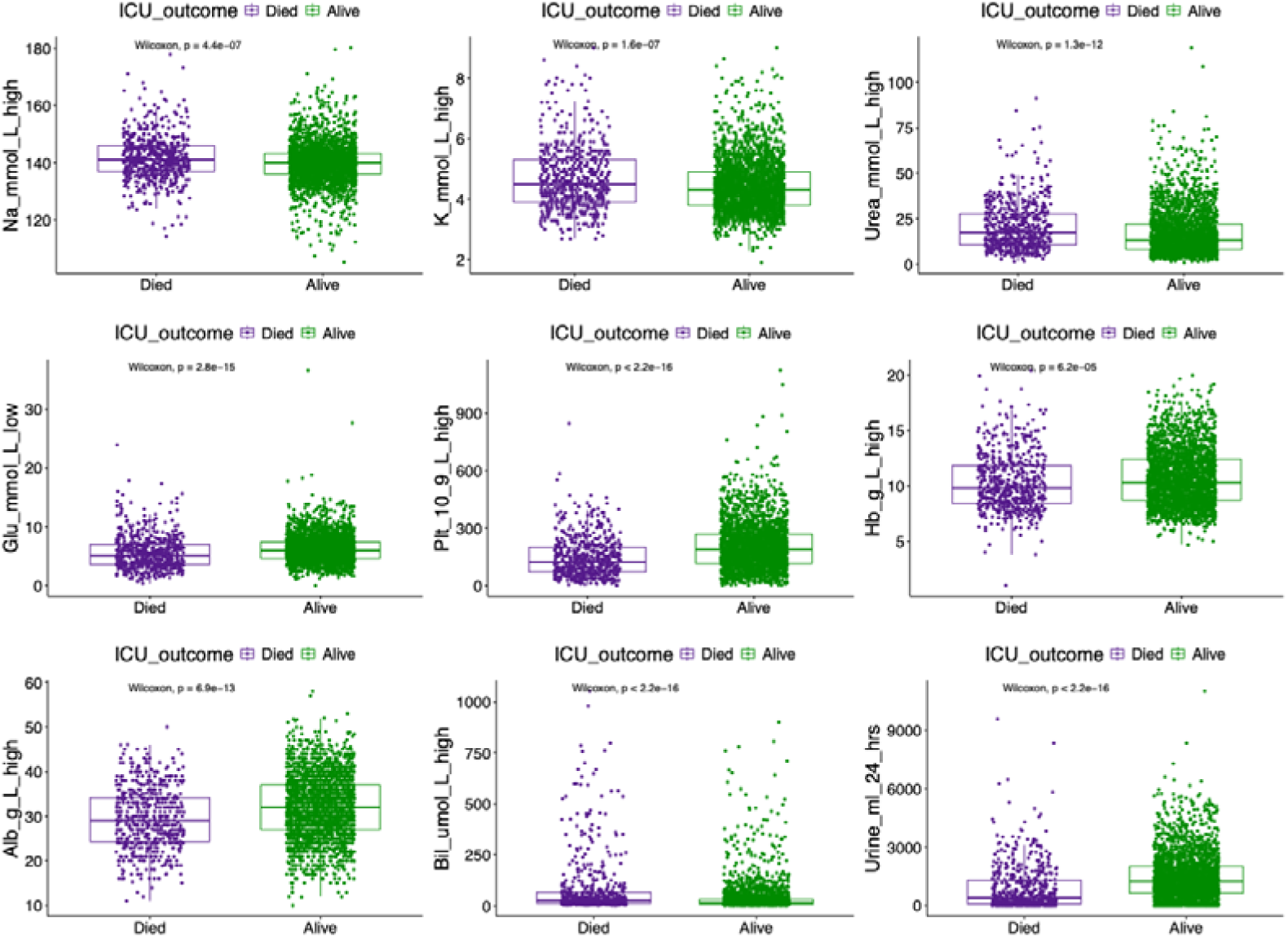
Association of ICU Mortality with Additional Clinical Parameters from Blood Tests

**Supplementary Figure 4.**
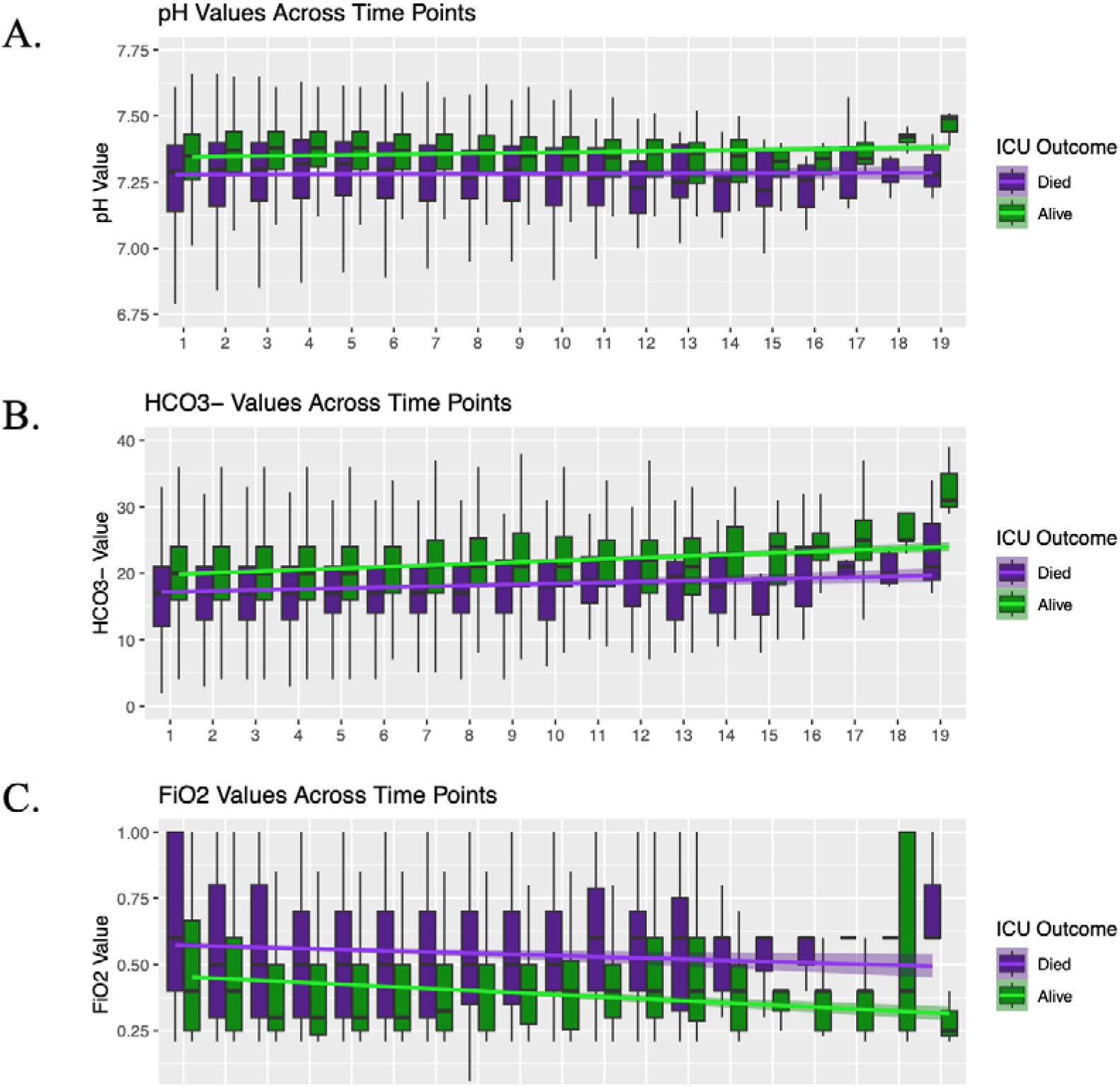
Time series trends and linear regression analyses by ICU mortality status.

**Supplementary Figure 5.**
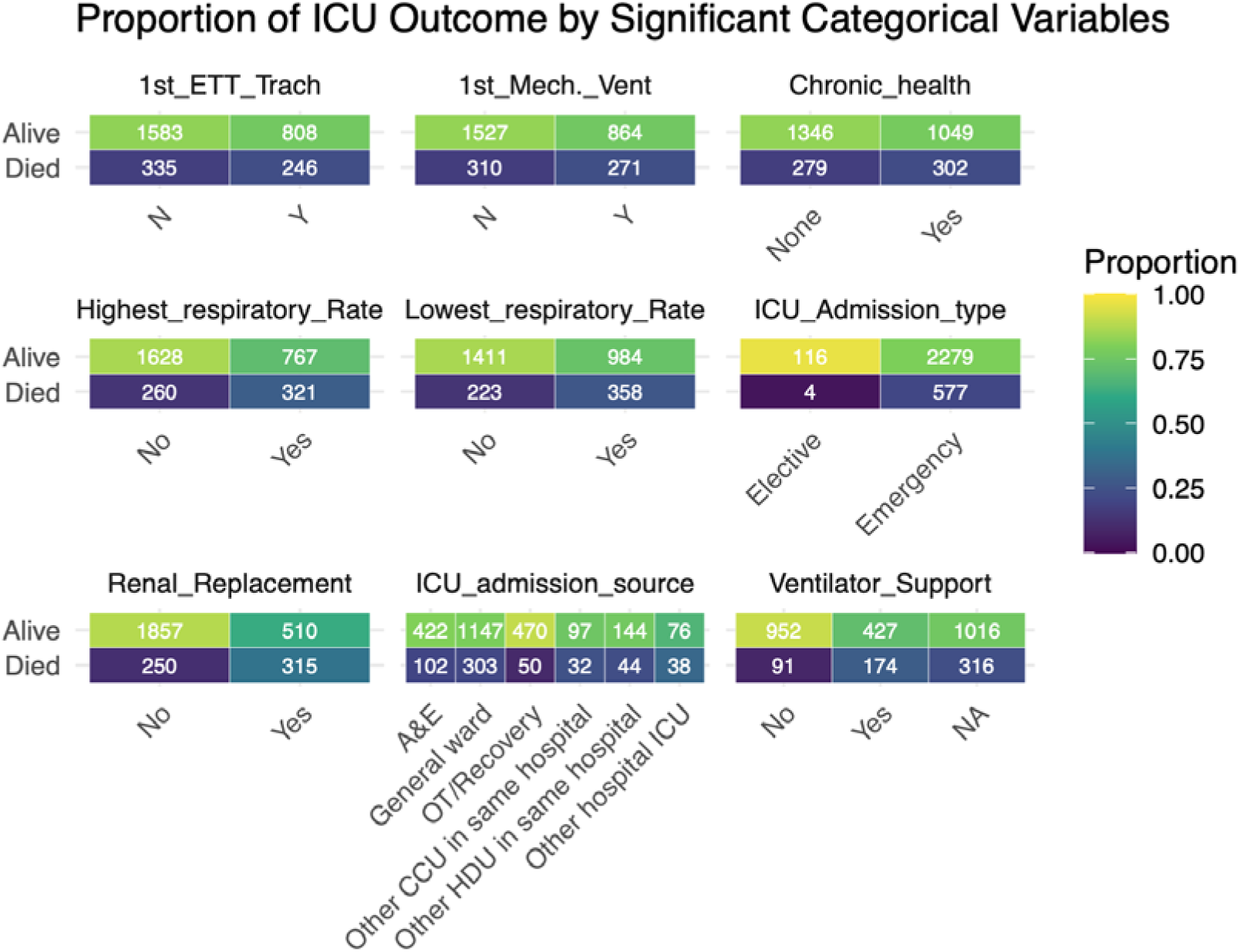
Proportion of ICU mortality stratified by significant categorical variables. Highest_respiratory_Rate: When measuring highest respiratory rate on ventilator (Y/N) Lowest_respiratory_Rate: When measuring lowest respiratory rate on ventilator (Y/N)

**Supplementary Figure 6.**
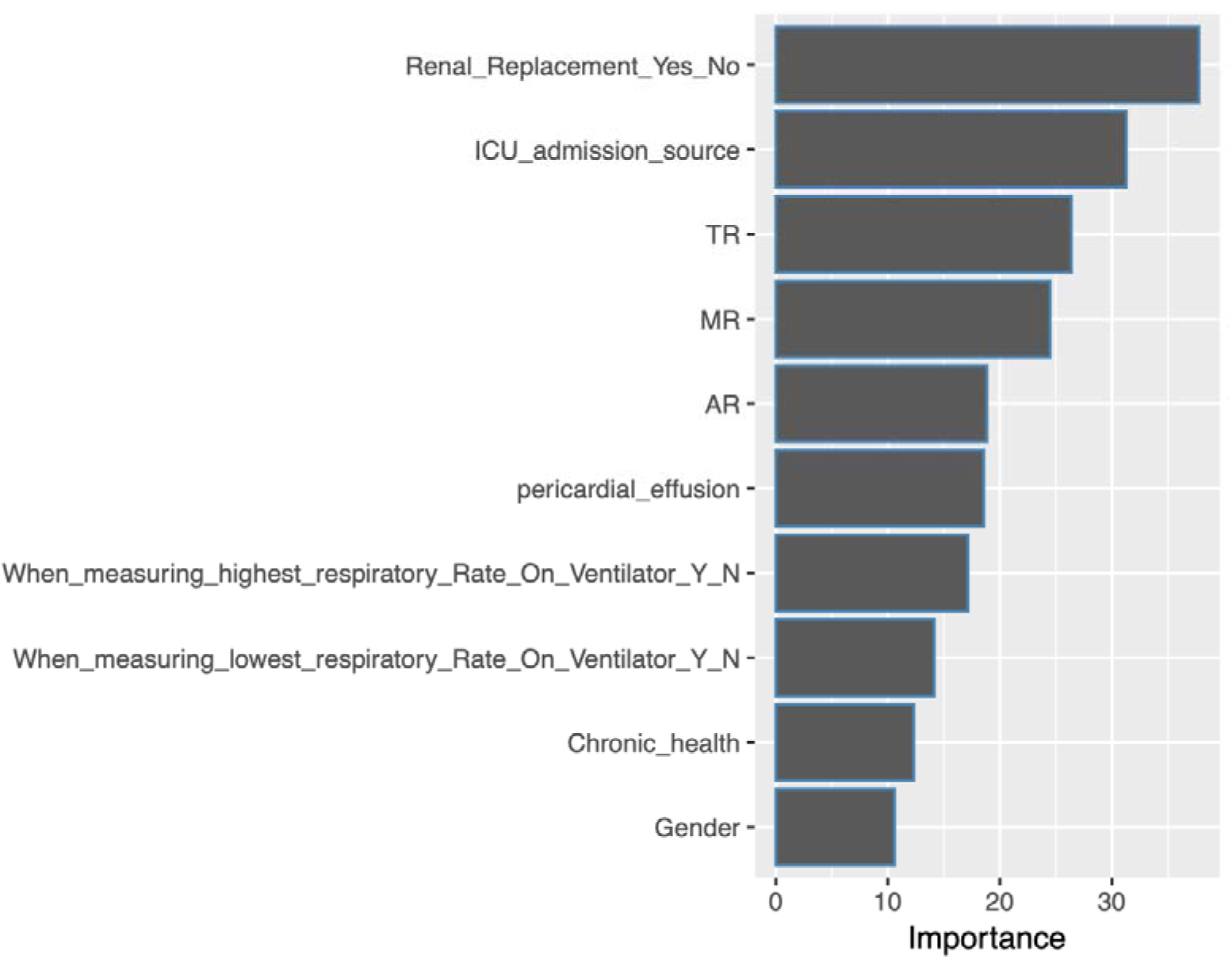
Importance scores of the top 10 categorical variables.

**Supplementary table 1:**
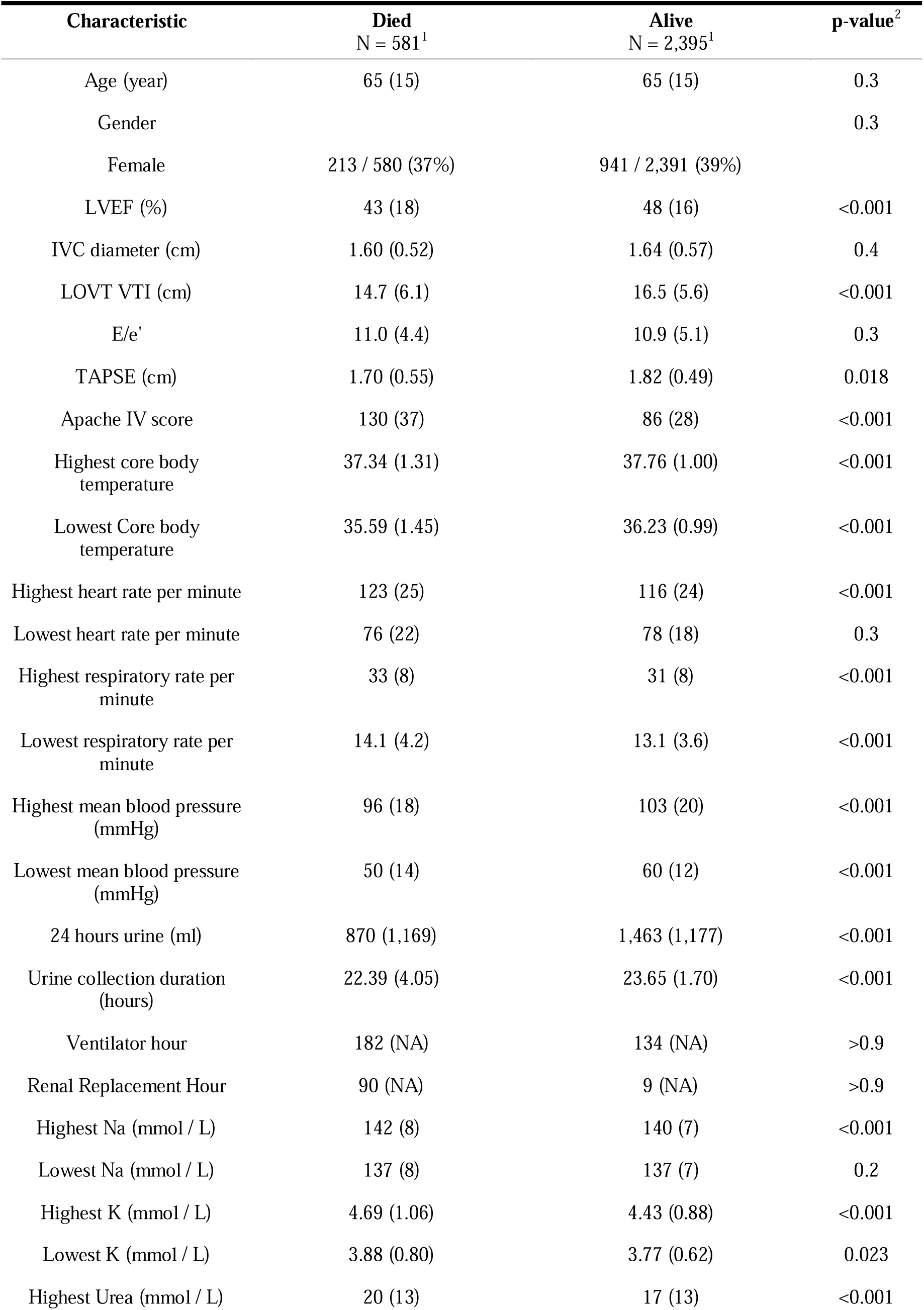
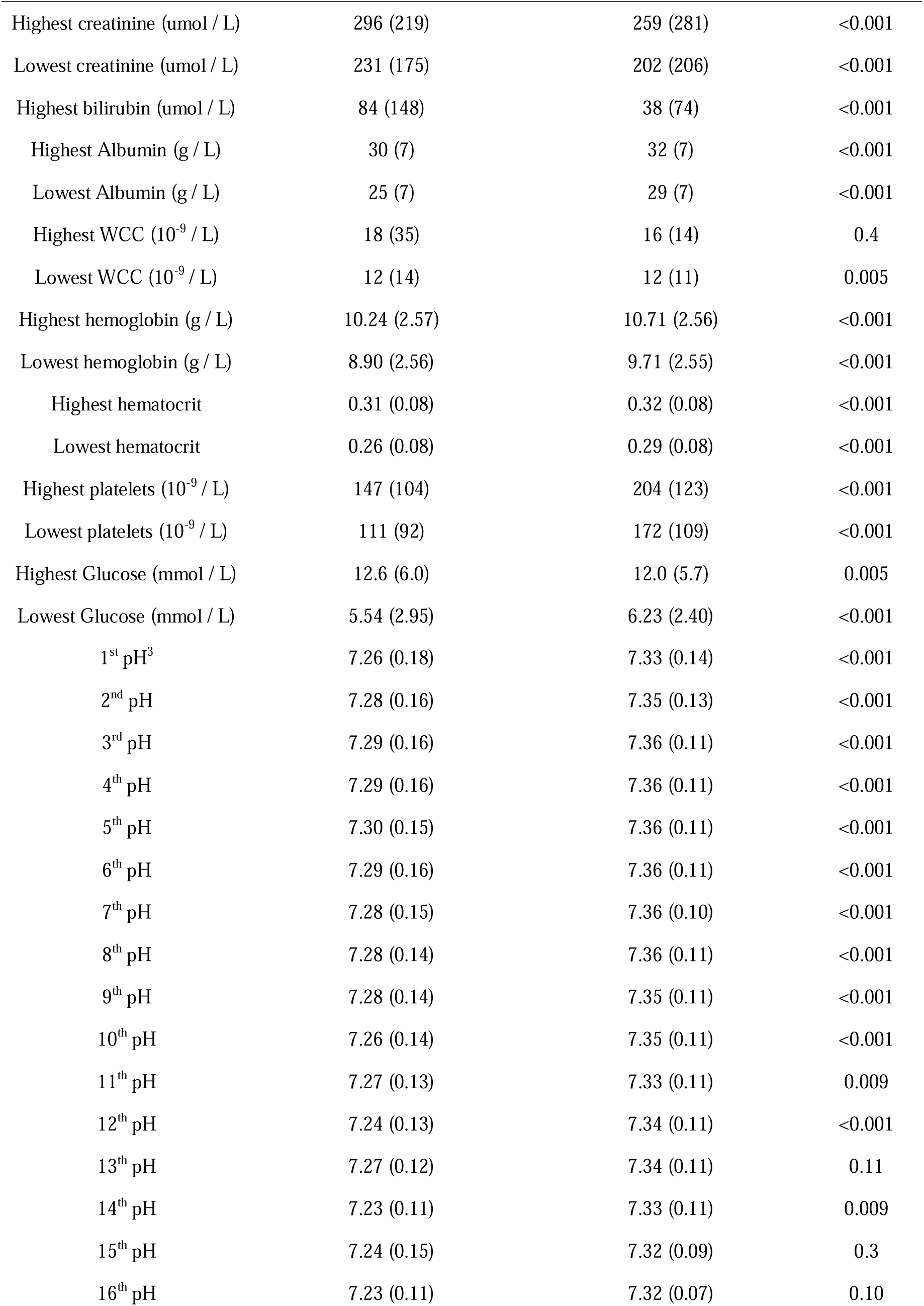
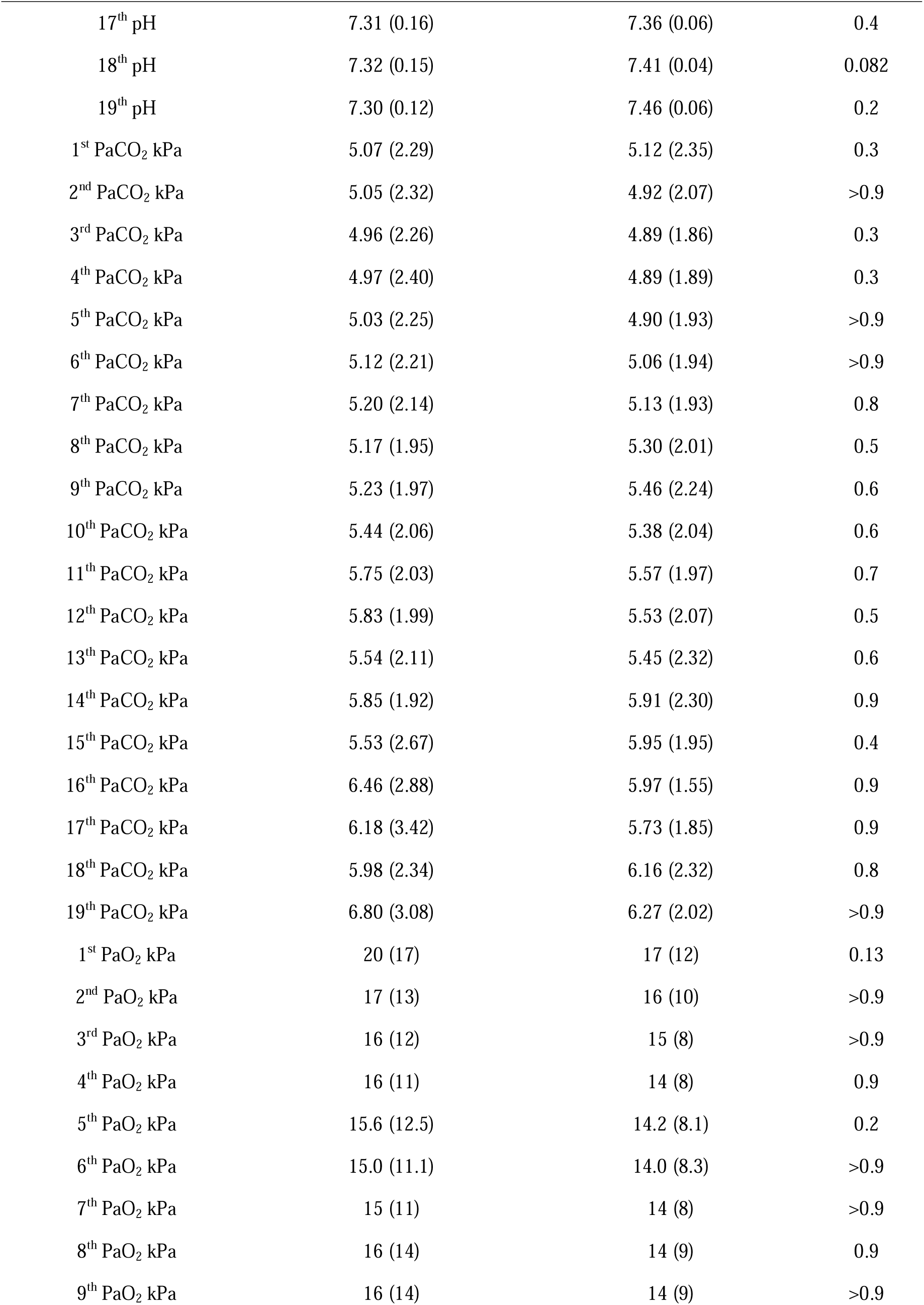
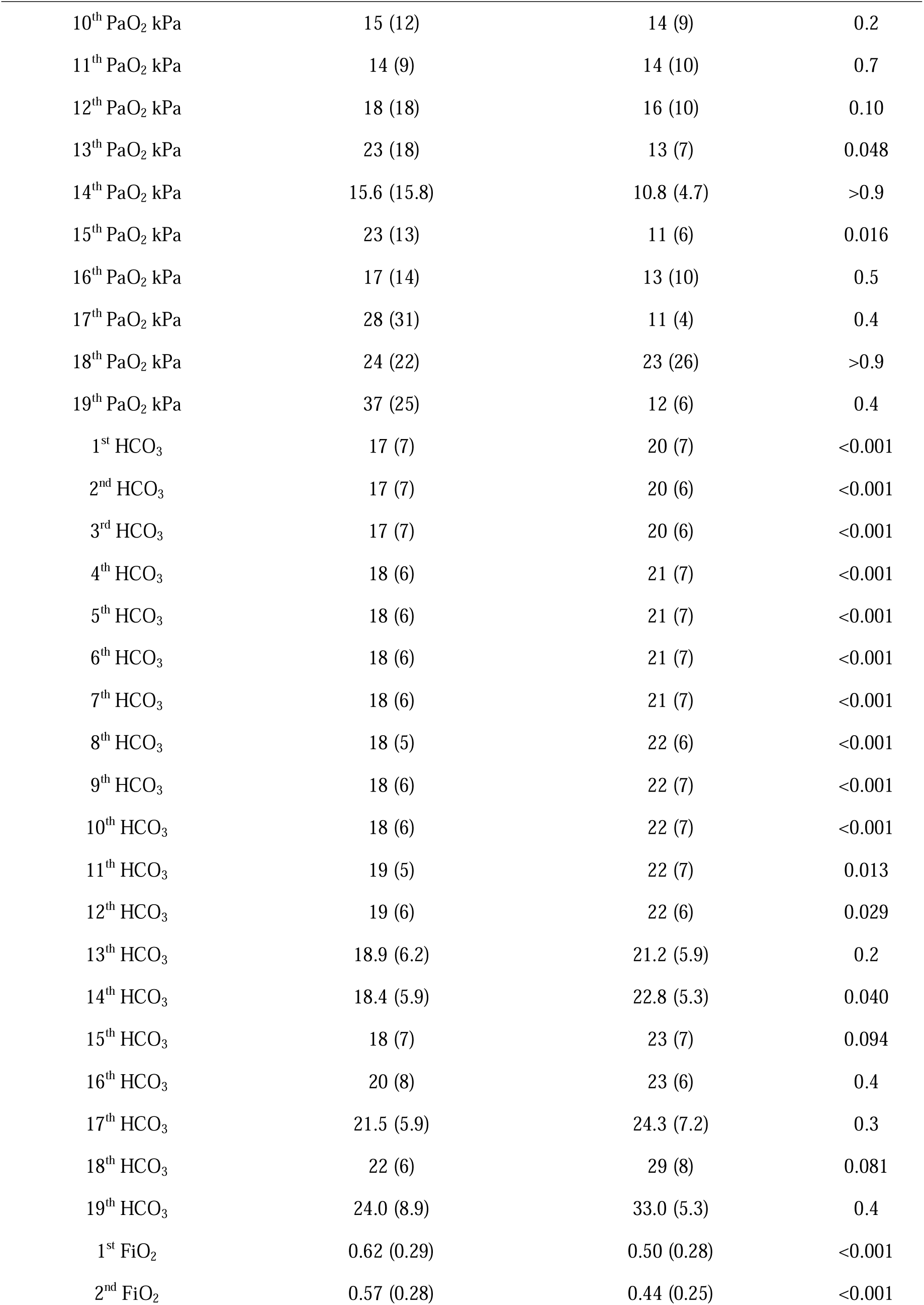
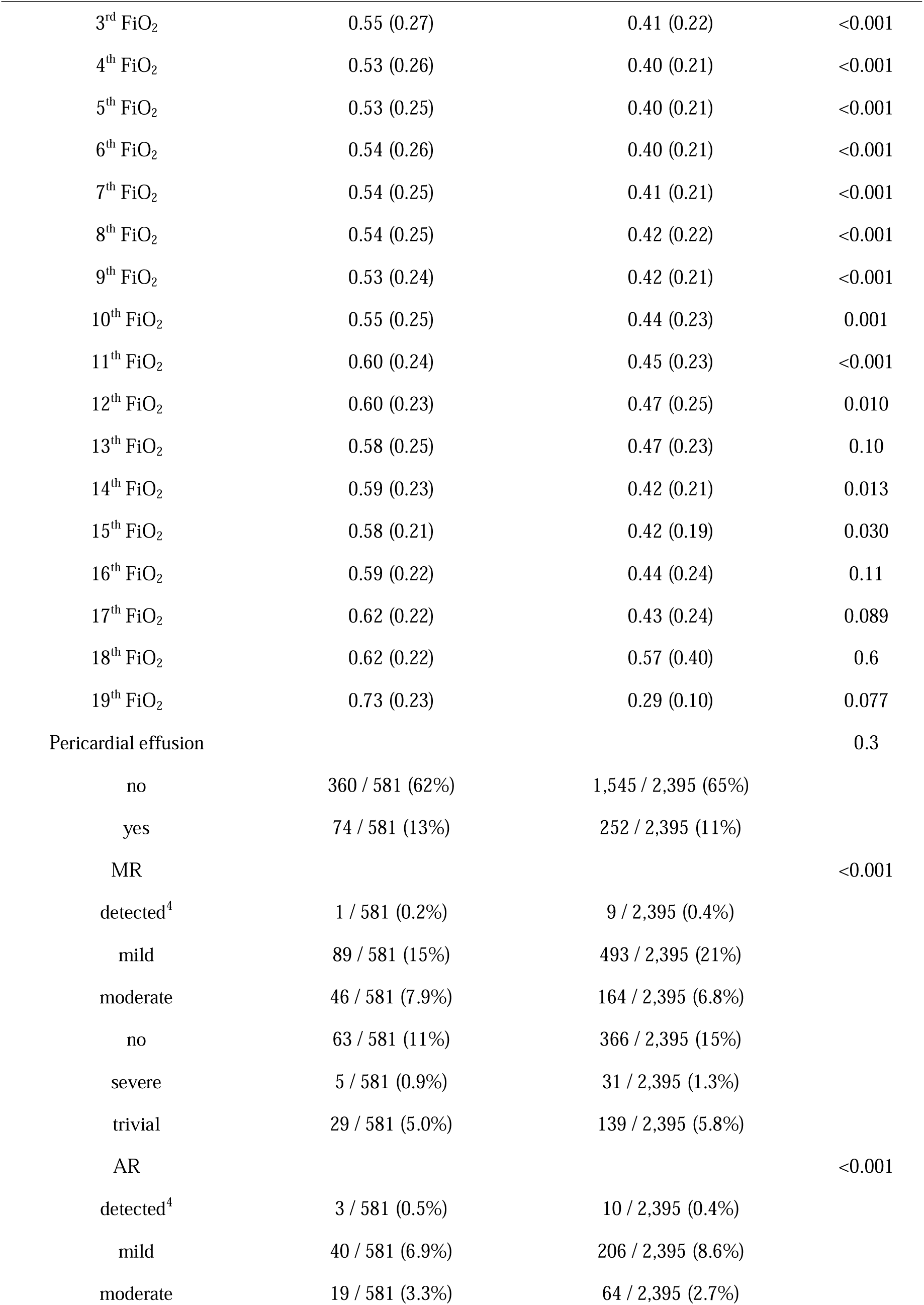
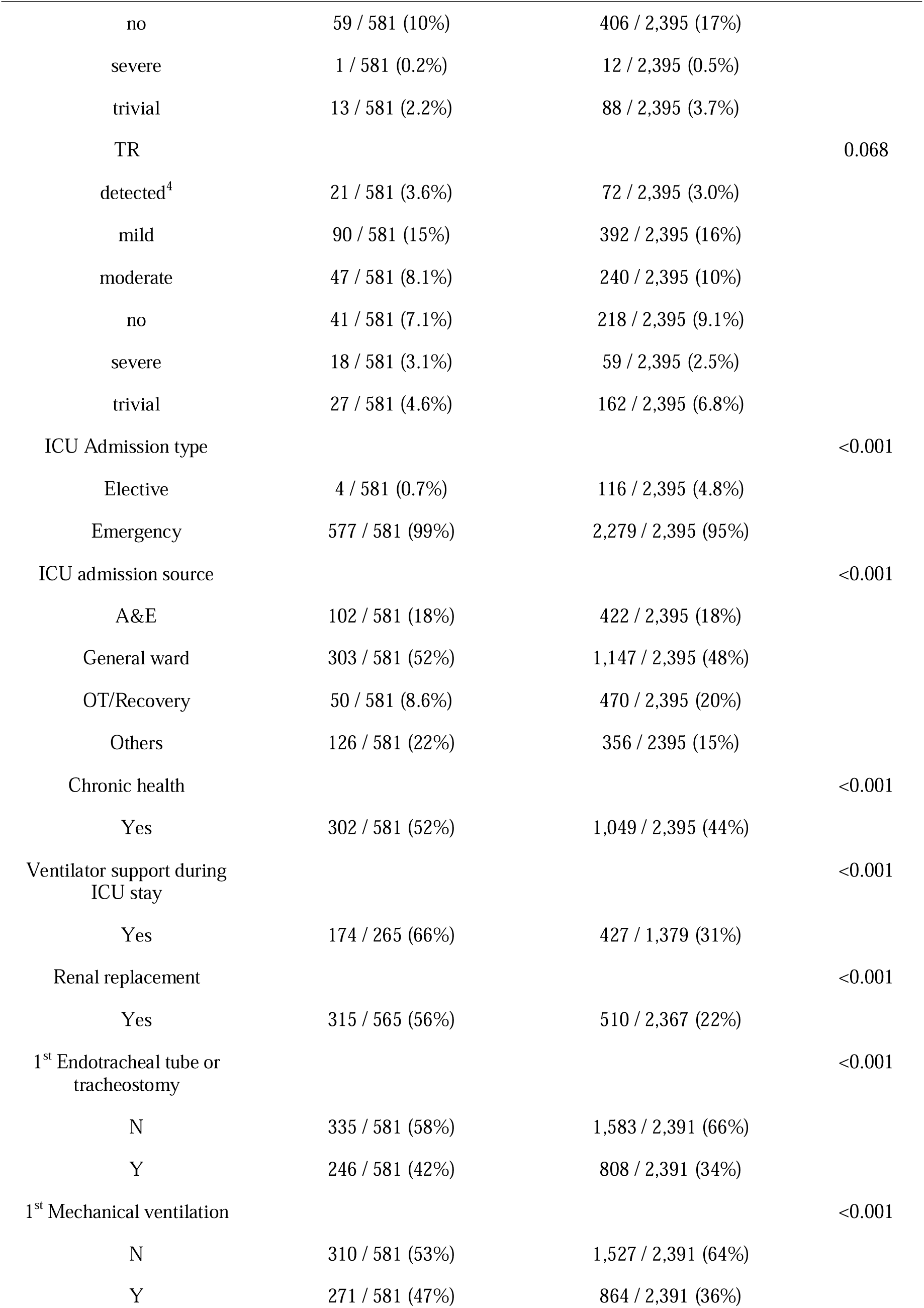
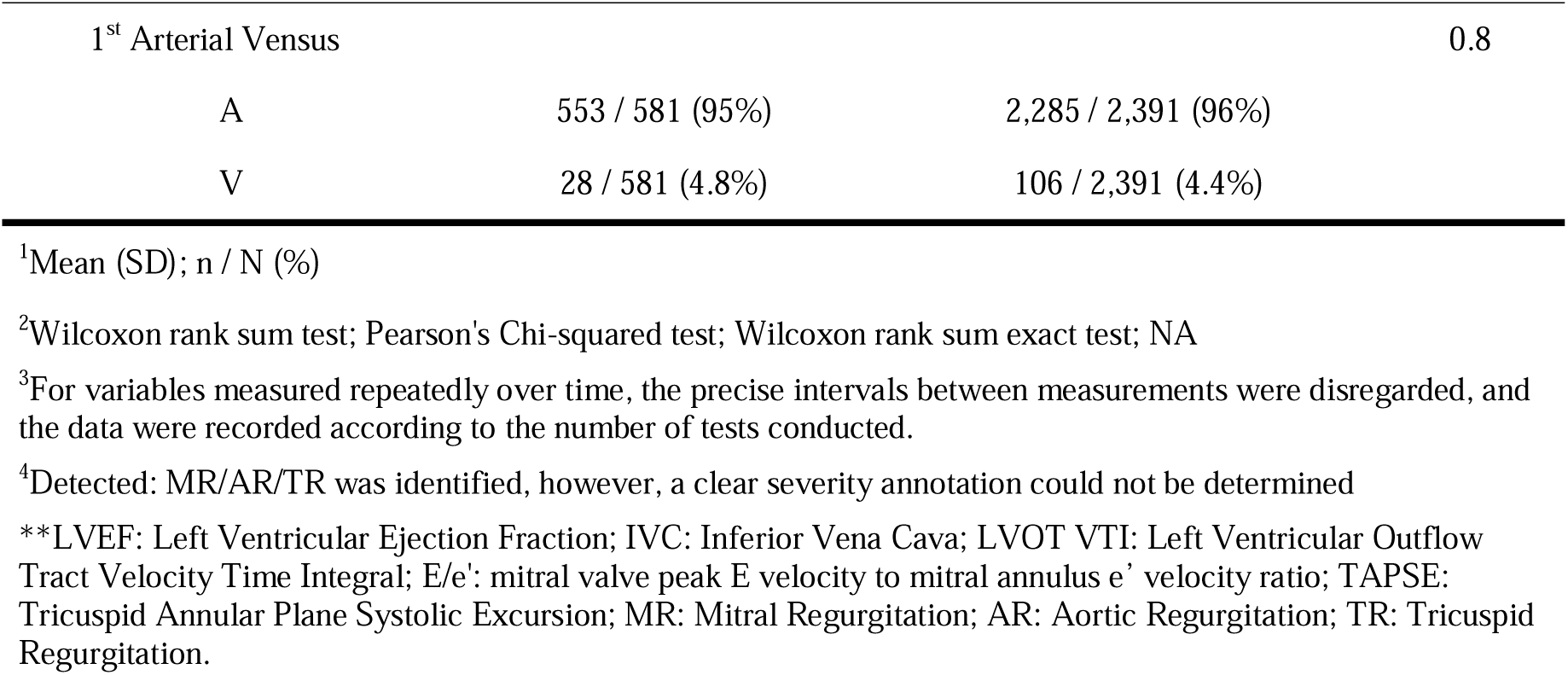
ICU clinical data.

**Supplementary table 2:**
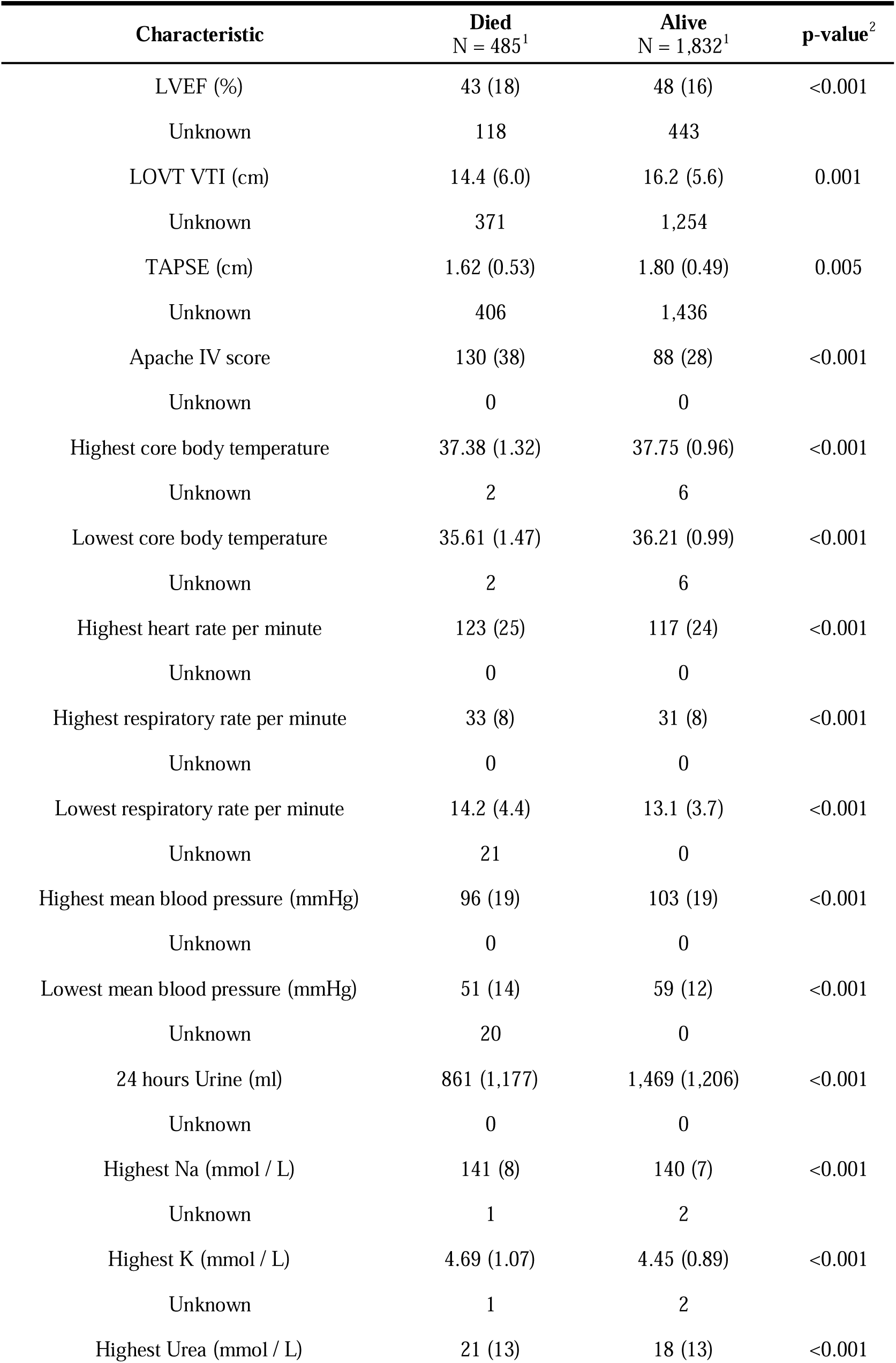
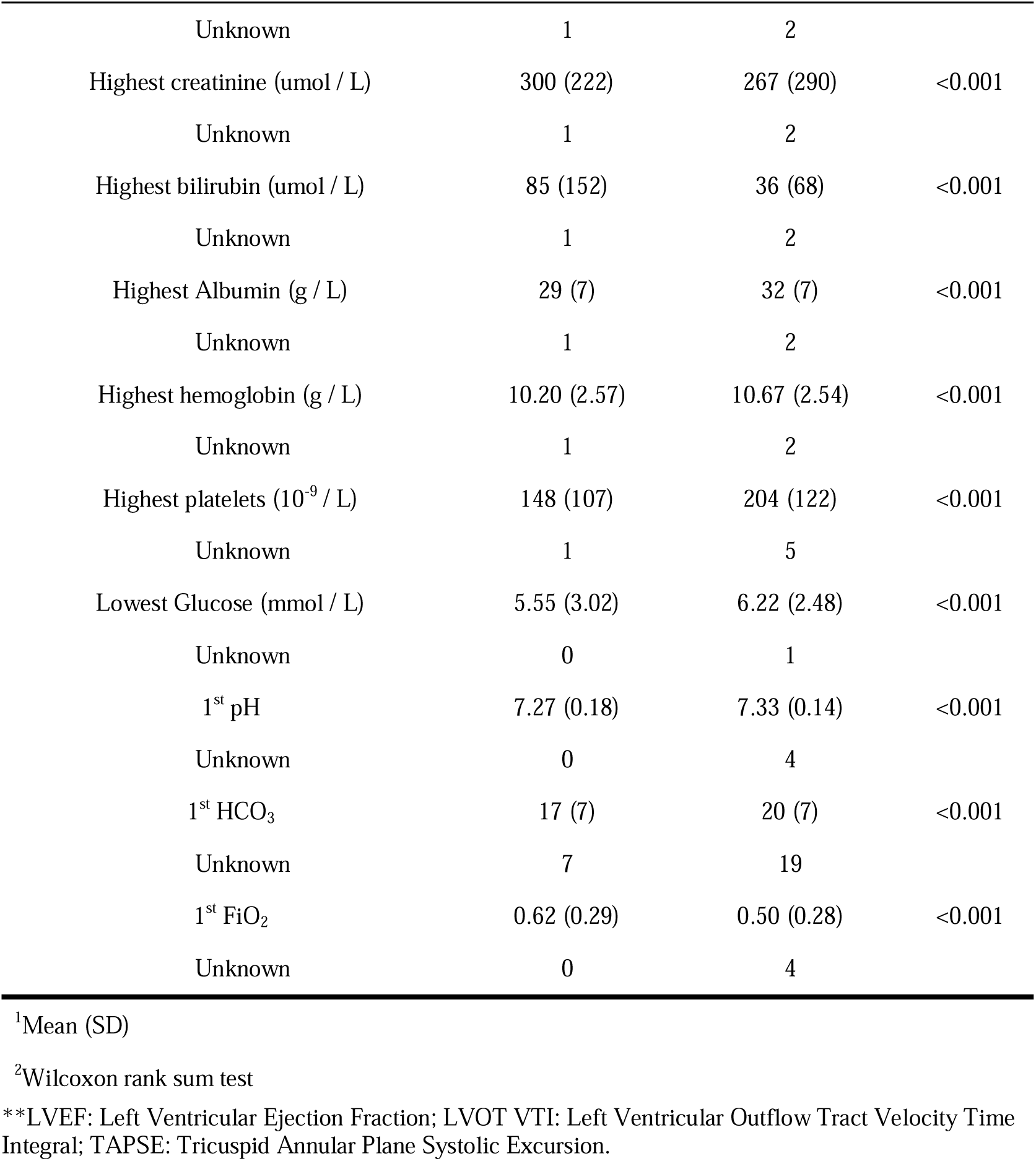
Continuous Variables Included in the Prediction Model for ICU Mortality.

| Characteristic | Died<br>N = 485<sup>1</sup> | Alive<br>N = 1,832<sup>1</sup> | p-value<sup>2</sup> |
| --- | --- | --- | --- |
| LVEF (%) | 43 (18) | 48 (16) | <0.001 |
| Unknown | 118 | 443 |  |
| LOVT VTI (cm) | 14.4 (6.0) | 16.2 (5.6) | 0.001 |
| Unknown | 371 | 1,254 |  |
| TAPSE (cm) | 1.62 (0.53) | 1.80 (0.49) | 0.005 |
| Unknown | 406 | 1,436 |  |
| Apache IV score | 130 (38) | 88 (28) | <0.001 |
| Unknown | 0 | 0 |  |
| Highest core body temperature | 37.38 (1.32) | 37.75 (0.96) | <0.001 |
| Unknown | 2 | 6 |  |
| Lowest core body temperature | 35.61 (1.47) | 36.21 (0.99) | <0.001 |
| Unknown | 2 | 6 |  |
| Highest heart rate per minute | 123 (25) | 117 (24) | <0.001 |
| Unknown | 0 | 0 |  |
| Highest respiratory rate per minute | 33 (8) | 31 (8) | <0.001 |
| Unknown | 0 | 0 |  |
| Lowest respiratory rate per minute | 14.2 (4.4) | 13.1 (3.7) | <0.001 |
| Unknown | 21 | 0 |  |
| Highest mean blood pressure (mmHg) | 96 (19) | 103 (19) | <0.001 |
| Unknown | 0 | 0 |  |
| Lowest mean blood pressure (mmHg) | 51 (14) | 59 (12) | <0.001 |
| Unknown | 20 | 0 |  |
| 24 hours Urine (ml) | 861 (1,177) | 1,469 (1,206) | <0.001 |
| Unknown | 0 | 0 |  |
| Highest Na (mmol / L) | 141 (8) | 140 (7) | <0.001 |
| Unknown | 1 | 2 |  |
| Highest K (mmol / L) | 4.69 (1.07) | 4.45 (0.89) | <0.001 |
| Unknown | 1 | 2 |  |
| Highest Urea (mmol / L) | 21 (13) | 18 (13) | <0.001 |
| Unknown | 1 | 2 |  |
| Highest creatinine (umol / L) | 300 (222) | 267 (290) | <0.001 |
| Unknown | 1 | 2 |  |
| Highest bilirubin (umol / L) | 85 (152) | 36 (68) | <0.001 |
| Unknown | 1 | 2 |  |
| Highest Albumin (g / L) | 29 (7) | 32 (7) | <0.001 |
| Unknown | 1 | 2 |  |
| Highest hemoglobin (g / L) | 10.20 (2.57) | 10.67 (2.54) | <0.001 |
| Unknown | 1 | 2 |  |
| Highest platelets (10<sup>-9</sup> / L) | 148 (107) | 204 (122) | <0.001 |
| Unknown | 1 | 5 |  |
| Lowest Glucose (mmol / L) | 5.55 (3.02) | 6.22 (2.48) | <0.001 |
| Unknown | 0 | 1 |  |
| 1<sup>st</sup> pH | 7.27 (0.18) | 7.33 (0.14) | <0.001 |
| Unknown | 0 | 4 |  |
| 1<sup>st</sup> HCO<sub>3</sub> | 17 (7) | 20 (7) | <0.001 |
| Unknown | 7 | 19 |  |
| 1<sup>st</sup> FiO<sub>2</sub> | 0.62 (0.29) | 0.50 (0.28) | <0.001 |
| Unknown | 0 | 4 |  |

<sup>1</sup>Mean (SD)
<sup>2</sup>Wilcoxon rank sum test
\*\*LVEF: Left Ventricular Ejection Fraction; LVOT VTI: Left Ventricular Outflow Tract Velocity Time Integral; TAPSE: Tricuspid Annular Plane Systolic Excursion.

